# Genetically determining individualized clinical reference ranges for the biomarker tryptase can limit unnecessary procedures and unmask myeloid neoplasms

**DOI:** 10.1101/2022.04.29.22274379

**Authors:** Jack Chovanec, Ilker Tunc, Jason Hughes, Joseph Halstead, Allyson Mateja, Yihui Liu, Michael P. O’Connell, Jiwon Kim, Young Hwan Park, Qinlu Wang, Quang Le, Mehdi Pirooznia, Neil N. Trivedi, Yun Bai, Yuzhi Yin, Amy P. Hsu, Josh McElwee, Sheryce Lassiter, Celeste Nelson, Judy Bandoh, Thomas DiMaggio, Julij Šelb, Matija Rijavec, Melody C. Carter, Hirsh D. Komarow, Vito Sabato, Joshua Steinberg, Kurt M. Hafer, Elizabeth Feuille, Christopher S. Hourigan, Justin Lack, Paneez Khoury, Irina Maric, Roberta Zanotti, Patrizia Bonadonna, Lawrence B. Schwartz, Joshua D. Milner, Sarah C. Glover, Didier G. Ebo, Peter Korošec, George H. Caughey, Erica H. Brittain., Ben Busby, Dean D. Metcalfe, Jonathan J. Lyons

## Abstract

Serum tryptase is a biomarker used to aid in the identification of certain myeloid neoplasms, most notably systemic mastocytosis, where baseline (BST) levels >20 ng/mL are a minor criterion for diagnosis. Whereas clonal myeloid neoplasms are rare, the common cause for elevated BST is the genetic trait hereditary alpha-tryptasemia (HαT) caused by increased germline *TPSAB1* copy number. To date, the precise structural variation and mechanism(s) underlying elevated BST in HαT and the general clinical utility of tryptase genotyping, remain undefined. Through cloning, long-read sequencing, and assembling of the human tryptase locus from an individual with HαT, and validating our findings *in vitro* and *in silico*, we demonstrate that BST elevations arise from over-expression of replicated *TPSAB1* loci encoding wild-type α-tryptase due to co-inheritance of a linked over-active promoter element. Modeling BST levels based upon *TPSAB1* replication number we generate new individualized clinical reference values for the upper limit of ‘normal’. Using this personalized laboratory medicine approach, we demonstrate the clinical utility of tryptase genotyping, finding that in the absence of HαT, BST levels >11.4 ng/mL frequently identify indolent clonal mast cell disease. Moreover, substantial BST elevations (e.g., >100 ng/mL) which would ordinarily prompt bone marrow biopsy, can result from *TPSAB1* replications alone and thus be within ‘normal’ limits for certain individuals with HαT.

## INTRODUCTION

The tryptase locus is structurally complex in humans (Table 1). Approximately two-thirds of people have α-tryptase, encoded at *TPSAB1* on one or both alleles, whereas everyone has β-tryptase encoded at one or both *TPSB2* alleles, as well as at non-α-tryptase encoding *TPSAB1* loci (1-3). Copy number gain and loss of tryptase gene sequences encoding both α- and β-tryptases have also been reported (4). However, the structures of such copy number variants (CNV) remain unknown. The most common CNVs observed among Western populations are *TPSAB1* gene replications encoding α-tryptase, a genetic trait known as hereditary alpha-tryptasemia (HαT) (5). HαT is inherited in an autosomal dominant pattern and has been shown to affect nearly 6% of the general population in the U.S., U.K., and E.U (6-9). *De novo* replications have not been reported. *TPSAB1* replications are associated with elevated basal serum tryptase (BST) levels of at least 2-3-fold higher than median BST levels of healthy individuals without HαT (10). While it has been hypothesized that additional *TPSAB1* copies identified in HαT are present within the tryptase locus at chromosome 16p13.3, and that over-expression of α-tryptase yields the disproportionate increases in BST levels seen relative to copy number (5), this has never been demonstrated. Moreover, since only a small region of *TPSAB1* has been probed to demonstrate gene replication (5, 11), it remains unknown whether extra-allelic copies of *TPSAB1* encode wild-type or novel sequences containing homology with α-tryptase at the probe site.

**Table 1.**
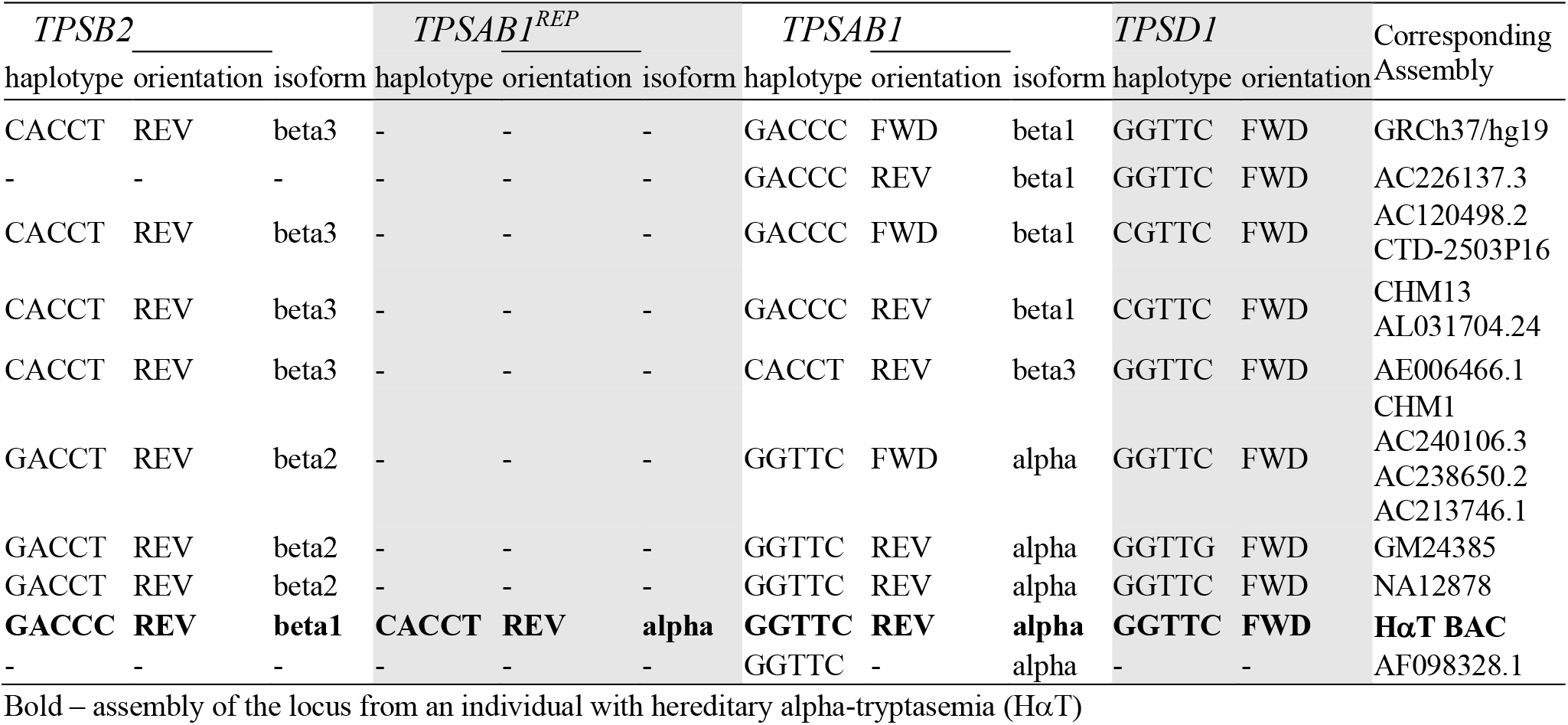
Assemblies of the human tryptase locus at 16p13.3.

Whereas HαT is the most common heritable cause for elevated BST, the clonal mast cell disorder systemic mastocytosis (SM) is a common acquired cause (10, 12), and BST levels are used routinely as a biomarker to screen for this disorder. Currently, the WHO criteria define a BST level >20 ng/mL as a minor criterion for the clinical diagnosis of SM (13) and this cut-off is frequently used in clinical decision making as an indication for bone marrow biopsy in symptomatic individuals (14). However, approximately one out of every four individuals with HαT has a BST level >20 ng/mL, representing an estimated 7.5 million people in the U.S. alone (4, 15). Because HαT has been shown to augment immediate hypersensitivity symptoms in a number of conditions (7, 8), many of these individuals are likely to undergo unwarranted invasive work-up for SM, including bone marrow biopsy, if tryptase genotype is not taken into account. Conversely, because HαT appears to account for most elevations in BST, observed elevations in patients who do not have HαT could be considerably enriched for other pathologies - most notably myeloid neoplasms - warranting workup when such elevations in BST may be modest (i.e., <20 ng/mL).

To address these questions, we cloned, long-read sequenced, and assembled the human tryptase locus containing a *TPSAB1* replication, finding the coding sequence to be wild-type α-tryptase. However, a series of unique proximal non-coding variants were also identified that distinguished replicated (α^DUP^-tryptase) from non-replicated (α^WT^-tryptase) sequences. An expanded DNA motif within the 5’-UTR was also linked to *TPSAB1* replication-associated variants and demonstrated increased *in vitro* promoter activity relative to the paralogous region in the non-replicated promoter. Using *in vitro* and *in silico* analyses of RNA sequences, we confirmed the relative over-expression of α^DUP^-tryptase sequences in primary basophils, cultured mast cells, and publicly available RNA-sequence datasets. Applying this knowledge, we generated a new genetic-based model for BST clinical reference ranges based upon *TPSAB1* replication number and examined the potential real-world impact of coupling tryptase genotyping with these newly defined values in the work-up of patients with clonal mast cell disorders. The potentially practice-changing implications for identification of indolent clonal mast cell disease, the diagnosis of SM, and the elimination of unnecessary bone marrow biopsies are discussed herein.

## RESULTS

### Increased TPSAB1 copy number occurs at the tryptase locus and encodes wild-type alpha-tryptase

A bacterial artificial chromosome (BAC) library was generated from an individual homozygous for *TPSAB1* duplications (genotype αα/β:αα/β) and a clone containing the complete human tryptase locus was identified. Using single molecule real time (SMRT) technology, we sequenced and subsequently assembled the locus *de novo* (Fig. 1a), allowing study of the structure and sequence of this variant-containing allele for the first time. As anticipated, the assembly displayed marked structural dissimilarity with the reference human genome (GRCh37/hg19) but did not contain novel sequence (Fig. S1, and online data repository). The additional *TPSAB1* copy encoding α-tryptase (α^DUP^) was located within the tryptase locus mapping to 16p13.3 between *TPSB2* and the non-replicated *TPSAB1* locus, as had been suggested based upon prior *in vitro* experiments (5). The coding sequence was identical to that of the adjacent wild-type α-tryptase sequence (α^WT^) (Table S4; online repository), and both sequences were in reverse orientation.

**Figure 1.**
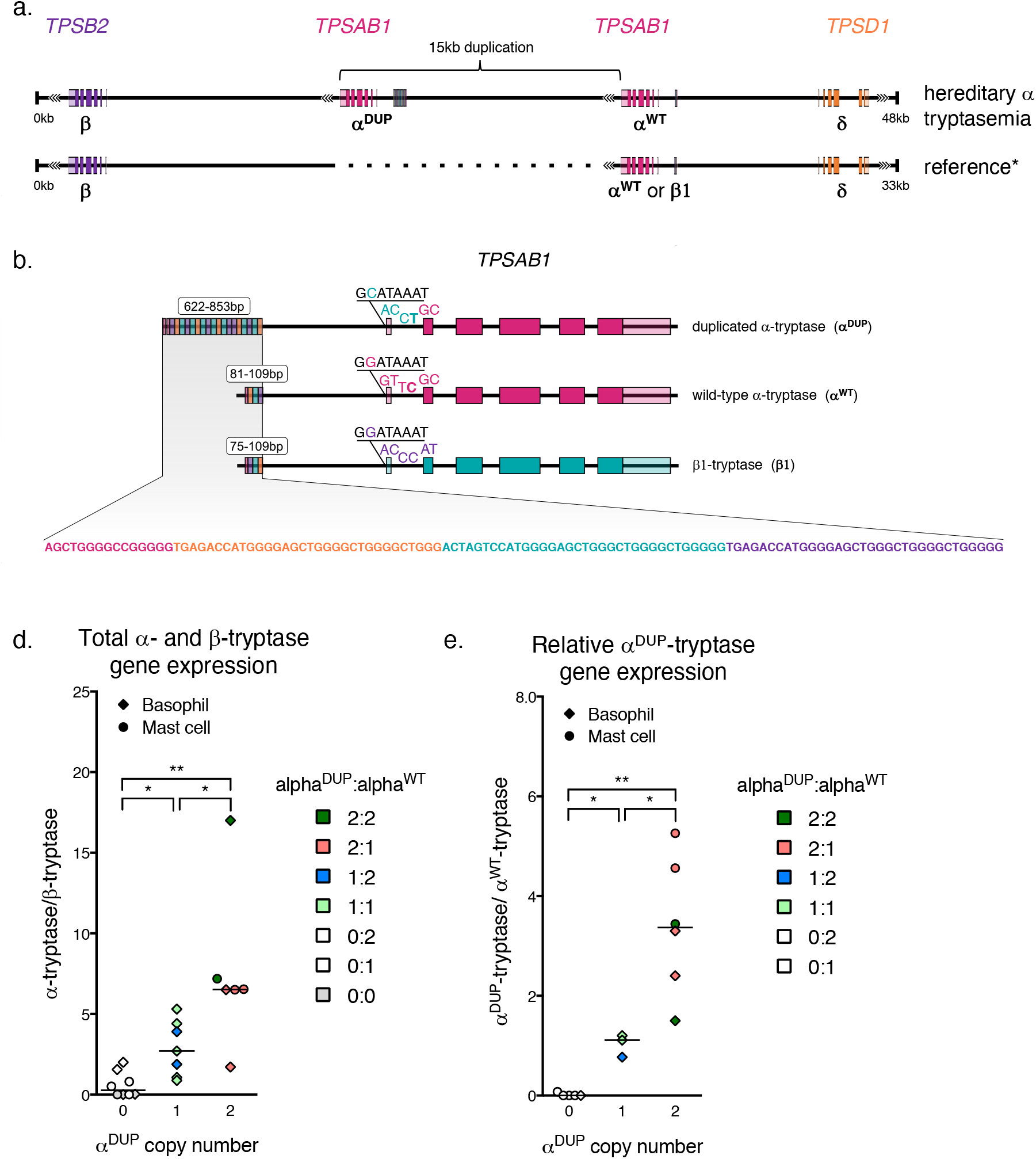
*TPSAB1* duplications occur at 16p13.3 and are linked to an expanded promoter associated with α^DUP^-tryptase over-expression. (**a)** Location and orientation of the 15-kB tandem duplication of *TPSAB1*. (**b)** Alignment of duplicated alpha- (α^DUP^), wild-type alpha (α^WT^), and beta1-tryptases with unique 5’-variants and size of promoter repeat regions indicated; repeat motif within promoter regions **(c)**. Relative total α- to β-tryptase gene expression **(d)** and α^DUP^-relative to α^WT^-tryptase gene expression **(e)** in ex-vivo basophils and cultured primary mast cells (right). *P<0.05; **P<0.005.

The sequence at *TPSB2* in our BAC clone was identified as β1-tryptase. While β1-tryptase at *TPSB2* is not common, it has been reported in approximately 5% of individuals screened from the HapMap cohort (16). Given that this prevalence was comparable to the prevalence of HαT reported in Western populations, we obtained the samples reported to contain β1-tryptase at *TPSB2* in order to determine whether they were all individuals with HαT. However, only 4 out of 15 of these individuals (27%) with β1-tryptase at *TPSB2* were found to have HαT. Thus, β1-tryptase at *TPSB2* is not exclusive to HαT.

### TPSAB1 replications encoding α^DUP^-tryptase are linked to an expanded promoter

Elevated BST levels among patients with HαT have been shown to result from increased constitutive release of pro-tryptases (5, 17). To determine whether this increase results from over-expression of α^DUP^-tryptase transcripts, we compared non-coding sequences at the two *TPSAB1* loci and identified an expanded (∼800 bp) repetitive DNA motif approximately 1 kilobase (kB) 5’ of the consensus transcription start site for α^DUP^-tryptase (Fig. 1b). This sequence was approximately 700 bp longer than the paralogous region of the α^WT^ promoter containing 27 repeated and 3 repeated sequences, respectively (Fig. 1c). Repeat regions at the same 5’ location were also observed in publicly available assemblies of α^WT^ sequences and confirmed by gel electrophoresis in amplified α^WT^-tryptase and α^DUP^-tryptase promoters from 9 unrelated individuals with HαT (Fig. S2). Subsequent cloning and Sanger sequencing of three additional α^WT^ and α^DUP^-tryptase promoters confirmed these relative differences (Fig. S3). Together these data demonstrate an expanded DNA repeat motif in the promoter of *TPSAB1* replications encoding α^DUP^-tryptase that is consistently co-inherited.

### Increased basal α^DUP^-tryptase expression is associated with elevated basal promoter activity

To quantify the functional outcome of the expanded promoter region linked to α^DUP^-tryptase sequences we next interrogated total α- and β-tryptase as well as α^WT^- and α^DUP^-tryptase transcript expression arising from *TPSAB1* and *TPSB2*. Cultured primary human mast cells and isolated basophils from individuals with HαT expressed higher levels of total α-tryptase transcripts, and over-expressed α^DUP^-tryptase sequences relative to other tryptase isoforms (Fig. 1d,e). Importantly, total α-tryptase gene expression levels were several orders of magnitude higher than those of β-tryptases, even when they were present in allelic balance (e.g., 2α,2β versus 3α,3β).

To determine if the expanded 5’ sequences uniquely associated with α^DUP^-tryptase contributed to the relative over-expression of α^DUP^-to α^WT^-tryptase, we next cloned the promoters containing these paralogous regions into fluorescent reporter plasmids using our BAC assembly and other SMRT-based *de novo* assemblies of the locus as sequence references (see methods for referenced assemblies). These constructs were sequence-verified and transfected into MonoMac-6 cells – an AML line that expresses tryptases (18, 19). The expanded α^DUP^-tryptase promoter demonstrated significantly greater basal activity relative to the α^WT^-tryptase promoter (Fig. 2a). This *in vitro* finding correlated directly with BST levels among individuals with HαT where the absolute number of *TPSAB1* replications was found to best correlate with BST level regardless of tryptase genotype derived from *TPSAB1* and *TPSB2* (Fig. 2b). Therefore, elevated BST in HαT results from over-expression of α^DUP^-tryptase at replicated *TPSAB1* loci linked to a promoter with increased basal activity.

**Figure 2.**
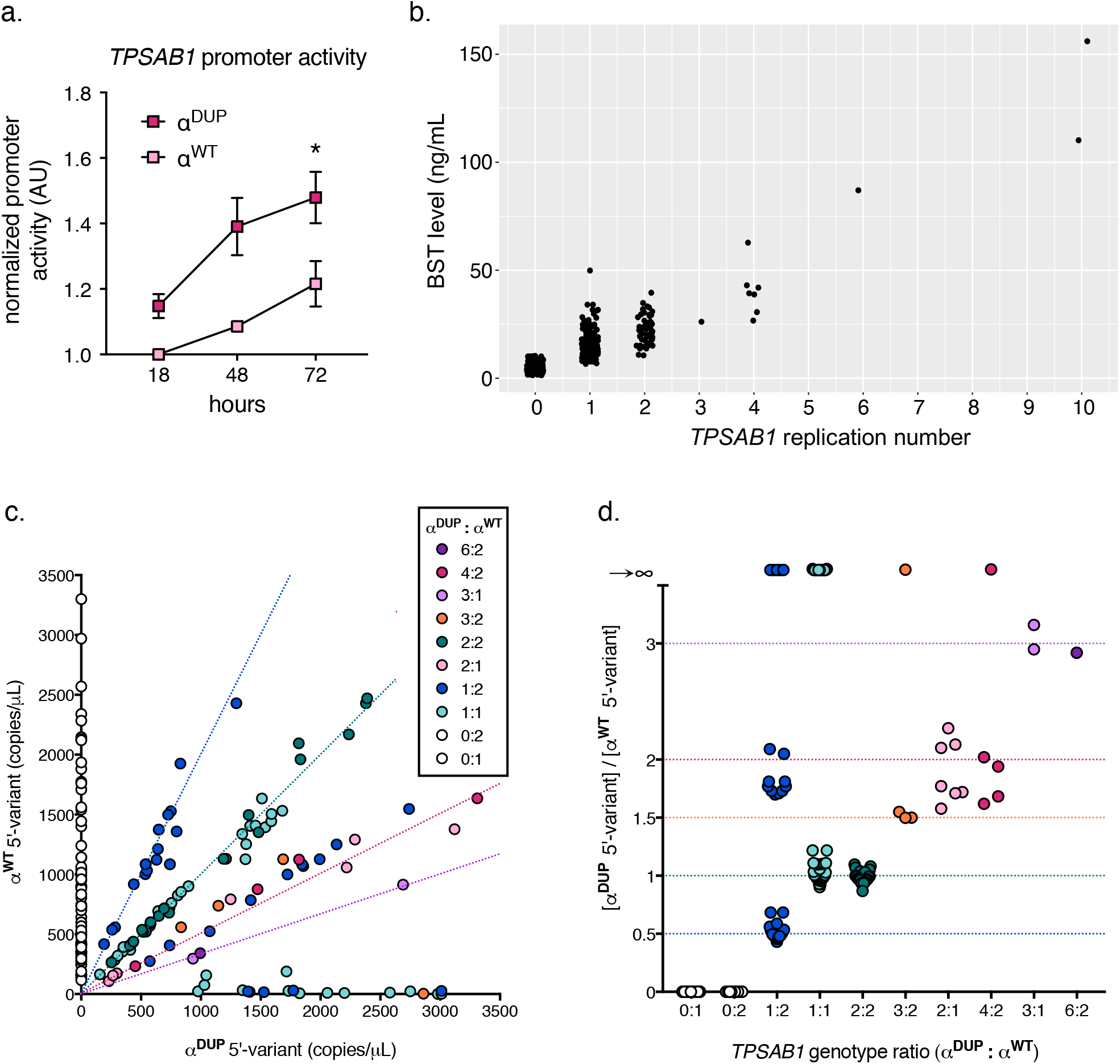
The expanded promoter at duplicated *TPSAB1* is conserved and has increased basal activity. **(a)** Normalized fluorescent-reporter activity in unstimulated MonoMac-6 cells transfected with promoters cloned from endogenous (α^WT^) or replicated (α^DUP^) *TPSAB1* loci. **(b)** Basal serum tryptase (BST) from individuals grouped by *TPSAB1* replication number. Relative allelic frequency **(c)** and ratios (d) of α^WT^-(y-axis) and α^DUP^-tryptase (x-axis) associated 5’-variants determined by droplet digital PCR; dashed lines indicate predicted α^WT^:α^DUP^ ratios by genotype. *P<0.05

### A unique haplotype demonstrates conservation of the identified over-active promoter at replicated TPSAB1 loci

To examine the generalizability of our finding of an expanded promoter linked to α^DUP^-tryptase at *TPSAB1*, we again examined proximal non-coding sequences associated with different tryptase isoforms and identified five unique 5’ variants – two substitutions in intron 1, two in the 5’ UTR, and one in the proximal promoter (Fig. 1b). Using these variants, we defined haplotypes, that when combined with coding sequences, uniquely distinguished α^DUP^-tryptase (CACCT) from α^WT^-tryptase (GGTTC), β1-tryptase (GACCC), and δ-tryptase (GGTTC) in our BAC assembly (Fig. 1b; see Table S1 for complete contextual sequences).

Next, we examined publicly available genome assemblies to determine if these haplotypes were conserved. Critically, *TPSAB1* and *TPSB2* have been shown to be in nearly complete linkage disequilibrium and are co-inherited in virtually all individuals such that major haplotypes exist and inheritance can be inferred (16, 20). The major tryptase isoforms encoded at *TPSAB1* are β1-tryptase followed by α^WT^-tryptase, and at *TPSB2* are β3-tryptase and β2-tryptase, when on β/β and α/β alleles respectively. As expected we observed GACCC linked to β1-tryptase at *TPSB2* on β1/β3 (β/β) alleles, and GGTTC linked to α^WT^-tryptase sequences at *TPSAB1* on α^WT^/β2 (α/β) alleles in all 13 queried assemblies as well as well as in the GRCh37/hg19 reference assembly (Table 1). Additional isoform-specific haplotypes at these two loci - GACCT linked to β2-tryptase at the *TPSB2* locus only when α^WT^/β2 (α/β) alleles were present and CACCT linked to β3-tryptases at the *TPSB2* locus only when β1/β3 (β/β) alleles were present - were also identified. Neither the α^DUP^-tryptase-specific promoter expansion nor the linked CACCT haplotype were observed in association with α^WT^-tryptase sequences in any of the seven assemblies containing α^WT^/β2 (α/β) (Table 1).

Using these haplotypes, we next re-examined genome sequence reads from 183 individuals without increased *TPSAB1* copy number, and from 32 affected individuals from 10 families with HαT (5). Employing a paired-read approach, we found that GACCC (β1-tryptase) and CACCT (β3-tryptase) were present in association with 98% (324/331) of individuals with β/β alleles. Moreover, the CACCT haplotype consistently segregated with the β/β haplotype in the 16 families without HαT in whom tryptase sequence inheritance could be examined. Whereas all α/β haplotypes (n = 6) were associated with the α^WT^-tryptase-linked GGTTC and β2-tryptase-linked GACCT (Table 2), the CACCT haplotype linked to α^DUP^-tryptase in our BAC clone, segregated universally with *TPSAB1* replications in all 32 individuals with HαT. The GACCC haplotype – linked to β1-tryptase most often present at *TPSAB1* but present at *TPSB2* in our BAC clone – was also found to be universally present among individuals with HαT and co-segregated with α^DUP^-tryptase containing alleles suggesting that β1-tryptase at *TPSB2* is frequently present on alleles containing *TPSAB1* replications, as seen in our BAC clone (Table 2). Therefore, these *in silico* findings suggest conservation of the unique promoter haplotypes we identified in our BAC assembly, and that over-expression of α^DUP^-tryptase could be a generalizable phenomenon.

**Table 2.** **Haplotypes identified in linkage with allelic tryptase genotypes.**

| *TPSB2* |  | *TPSAB1<sup>DUP</sup>* |  | *TPSAB1* |  | *TPSD1* | Allele count | Allelic genotype | Haplotype Frequency (by genotype) |
| --- | --- | --- | --- | --- | --- | --- | --- | --- | --- |
| haplotype | isoform | haplotype | isoform | haplotype | isoform | haplotype |  |  |  |
| CACCT | beta3 | - | - | GACCC | beta1 | GGTTC | 172 | β/β | 52% |
| CACCT | beta3 | - | - | GACCC | beta1 | CGTTC | 152 | β/β | 46% |
| GACCT | beta2 | - | - | GACCC | beta1 | GGTTC | 7 | β/β | 2% |
| GACCT | beta2 | - | - | GGTTC | alpha | GGTTC | 65 | α/β | 96% |
| GACCT | beta2 | - | - | GGTTC | alpha | CGTTC | 3 | α/β | 4% |
| GACCC | beta1 | CACCT | alpha | GGTTC | alpha | GGTTC | 15 | αα/β | 63% |
| GACCC | beta1 | CACCT | alpha | - - - - T | alpha | GGTTC | 5 | αα/β | 21% |
| GACCC | beta1 | CACCT | alpha | GGTT (-6delC)C* | alpha | GGTTC | 2 | αα/β | 8% |
| GACCC | beta1 | CACCT | alpha | GGTTT | alpha | GGTTC | 2 | αα/β | 8% |

\*A single cytosine (C) base deletion was identified 6 bases prior (-6) to the terminal C of the haplotype

To confirm these findings at scale *in vitro*, we had to overcome the repetitive and GC-rich nature of the region. To do so we developed a ddPCR assay capable of detecting the most proximal variant which distinguished the α^DUP^-tryptase promoter haplotype (CACC**<u>T</u>**) from that of α^WT^-tryptase (GGTT**<u>C</u>**) (see Fig. S4 for description and representative data). The corresponding variant listed in the reference genome (GRCh37/hg19 16:1290818C>T) has only been reported in association with *TPSB2* – ostensibly linked to dominant isoform β3-tryptase at this locus. However, the reverse primer in our assay was designed to only hybridize with α-tryptase sequences.

The CACC**<u>T</u>** variant was confirmed to be universally present in linkage with α^DUP^-tryptase in 120 affected individuals with increased *TPSAB1* copy number from 101 families. Conversely, CACC**<u>T</u>** was never found in linkage with α^WT^-tryptase in 81 unaffected individuals from 69 families who did not have increased *TPSAB1* copy number but carried one or two α^WT^-tryptase-encoding copies (Fig. 2c). Furthermore, the ratios of identified promoter copy number confirmed that all 166 HαT-associated α^DUP^-tryptase sequences on 156 alleles were linked to the C>T variant (Fig. 2d) and by inference, the expanded promoter.

Interestingly, in 30/120 individuals with HαT, α^WT^-tryptase was not linked to GGTT**<u>C</u>**; 10 of these α^WT^-tryptase promoters appeared to contain the C>T missense, and the remaining 19 alleles may have had this or another unknown missense variant or indel, the latter of which may have resulted in failed probe or primer hybridization. Importantly, our *in silico* analysis identified a GGTTT haplotype linked to α^WT^-tryptase in one family with HαT. In a second family a single base-pair deletion in the -6 position from C>T was observed, which we have labelled GGTT(−6delC)C was also identified (Table 2). This deletion interfered with probe hybridization in our assay potentially accounting for some of the 19 α^WT^-tryptase containing alleles with ambiguous promoter linkage. The α^WT^-tryptase promoter from these samples also failed to amplify and was not successfully cloned, suggesting yet another possible α^WT^-tryptase promoter may exist. Indeed, public database interrogation yielded at least six different α-tryptase sequences exhibiting a high degree of homology (Table S4). Regardless, α^WT^-tryptase containing alleles with undefined promoter haplotypes, or even those linked to the C>T intronic variant, were not associated BST differences *in vivo* (Fig. S5), indicating that these undefined promoters are not associated with differences in gene expression at linked α^WT^-tryptase encoding *TPSAB1* loci. Collectively, these data support conservation of a unique haplotype linked to an over-active promoter element at replicated *TPSAB1* loci that is associated with over-expression of α-tryptase by mast cells and basophils.

### α^DUP^-Tryptase transcripts are over-expressed in public datasets

Using α- AND β-tryptase isoform-specific coding variants and the uniquely linked proximal non-coding haplotypes, a 39-base consensus sequence was defined that contained four variants which could distinguish α^WT^-, α^DUP^-, and β-tryptase transcripts; given the proximity of the first variant to the transcriptional start site, only the distal three variants were used for read alignment (Fig. 1b, Table S2). Querying 58 non-disease-associated transcriptome datasets, 863/4160 samples were identified with exact transcript matches to ≥1 of the three defined 39-bp consensus sequences. α^DUP^-Tryptase was observed in 3% (25/863) and α^WT^-tryptase transcripts were identified in 34% of individuals (289/863) (Table S5). Because linked meta-data were limited, the race and ethnicities of these samples are unknown. While Caucasians are likely to be over-represented in these samples, other ethnicities and racial groups are likely included which may explain the lower-than-expected prevalence of HαT (3% v. 5.6%).

Using the same method, we next examined the expression levels of α^WT^-, α^DUP^-, and β-tryptase transcripts in these and other public datasets. This analysis confirmed over-expression of both α^WT^-, α^DUP^-tryptase transcripts relative to β-tryptase (Fig. 3a). While over-expression of α^WT^-tryptase was also observed in our primary gene expression data, it is possible that some δ-tryptase reads could align to the α^WT^-tryptase consensus sequence, due to haplotype sequence homology, but not to α^DUP^-tryptase where the linked haplotype is not observed in linkage with δ-tryptase. Therefore, misalignment of δ-tryptase reads could lead to a potential over-estimation of α^WT^-tryptase expression levels. To exclude this possibility, we examined BST levels – which only measure total secreted α- and β-tryptases – among individuals without HαT. As reported (21), we saw a modest but significant positive correlation between BST levels and increasing α^WT^-tryptase copy number at *TPSAB1* (Fig. 3b). Thus, consistent with the RNA sequence read alignment results, α^WT^ does appear to be modestly over-expressed relative to β-tryptases. Because of the variable levels of total tryptase transcript in any given sample, we next employed regression analysis to quantify the relative expression of α^DUP^-tryptase to α^WT^-tryptase more precisely. Consistent with all our prior data, α^DUP^-tryptase was found to be over-expressed relative to α^WT^-tryptase by a factor of 5.2-fold (95% CI 4.864 to 5.499) in these datasets (R^2^=0.8, P<0.0001) (Fig. 3c). Thus, *in silico, in vitro*, and *in vivo* findings collectively indicate that elevated BST in HαT results from basal over-expression of wild-type α-tryptase at replicated *TPSAB1* loci linked to co-inherited over-active promoter elements.

**Figure 3.**
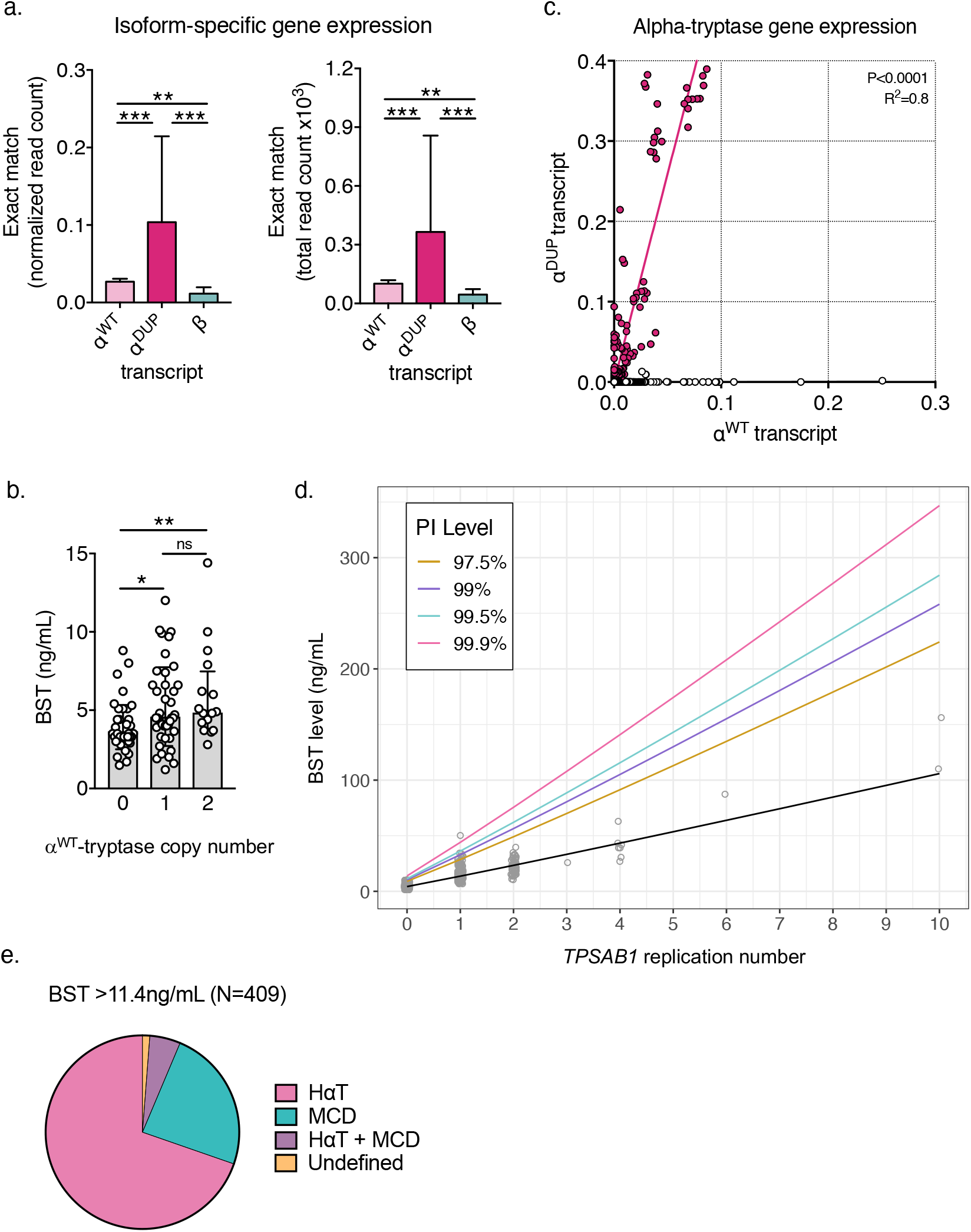
Modeling serum tryptase levels based upon genotype improves clinical utility. **(a)** Normalized (left) and total (right) read counts for reads aligning exactly to the 39-bp consensus sequences that identify β-, α^WT^- and α^DUP^-tryptase. **(b)** BST levels among individuals with conserved 4n tryptase copy number (combined from *TPSAB1* and *TPSB2*). **(c)** Regression analysis of relative expression levels of α^DUP^ (y-axis) and α^WT^ (x-axis) tryptase transcripts. (d) Prediction intervals (PI) for BST levels based upon *TPSAB1* replication number. (e) Prevalence of HαT, clonal mast cell disease (MCD), and those without either, among individuals referred with BST levels above the predicted upper limit of normal (>11.4 ng/mL). *P<0.01; **P<0.005; ***P<0.0001

### Modeling BST levels based upon TPSAB1 replication number redefines clinical reference ranges

Using this knowledge, we created a data-driven model from 204 individuals with normal *TPSAB1* copy number and 309 individuals with HαT in whom there was no clinical evidence of clonal mast cell disease (MCD), to generate prediction intervals with upper limits for predicted BST levels based upon *TPSAB1* replication number (Fig. 3d) and created on online application for clinical use: **<u>B</u>**asal **<u>S</u>**erum **<u>T</u>**ryptase **<u>C</u>**linical cut-off **<u>A</u>**ssigned by **<u>L</u>**ocus **<u>C</u>**opy number of **<u>U</u>**TR-**<u>L</u>**inked element and **<u>A</u>**ssociated **<u>*T*</u>***PSAB1* **<u>E</u>**ncoded **<u>R</u>**eplication (BST CALCULATER) available at: https://bst-calculater.niaid.nih.gov/. These thresholds can be used to determine where a bone marrow or further clinical evaluation may be needed for individuals with or without HαT when another clinical indication for such evaluation may be lacking.

This model not only redefined clinically meaningful upper limits for BST levels among individuals with HαT, but it also for the first time defines a clinically actionable upper limit among individuals without HαT as 11.4 ng/mL (Table 3). Remarkably, this has been the upper limit of normal for BST commonly used in most clinical laboratories base. Moreover, based upon the data, this cut-off of 11.4 ng/mL would appear to be valid in defining a clinically abnormal elevated BST when considering the diagnosis of clonal mast cell disease - as opposed to the currently accepted 20 ng/mL - provided that HαT has been ruled out by tryptase genotyping.

**Table 3.** **Measured and predicted BST levels based upon *TPSAB1* replication number encoding α-tryptase.**

| *TPSAB1*<br>replication<br>number | Primary data |  |  | Modeled reference<br>values |  |
| --- | --- | --- | --- | --- | --- |
|  | N | Median<br>BST | SD BST | Fit | Upper 99.5%<br>PI |
| 0 | 204 | 4.15 | 2.12 | 4.2831 | 11.3879 |
| 1 | 247 | 13.60 | 5.55 | 13.6363 | 36.2216 |
| 2 | 52 | 22.45 | 6.40 | 23.3642 | 62.1517 |
| 3 | 1 | 26.00 | NA | 33.3110 | 88.7541 |
| 4 | 6 | 40.50 | 12.61 | 43.4148 | 115.8535 |
| 5 | - | - | - | 53.6420 | 143.3508 |
| 6 | 1 | 87.00 | NA | 63.9708 | 171.1816 |
| 7 | - | - | - | 74.3862 | 199.3007 |
| 8 | - | - | - | 84.8774 | 227.6741 |
| 9 | - | - | - | 95.4357 | 256.2756 |
| 10 | 2 | 133.00 | 32.53 | 106.0544 | 285.0841 |

BST - basal serum tryptase; SD - standard deviation; PI - prediction interval

This model fit the primary data well (R^2^=0.76, P<0.001) with only one individual being identified in the entire primary dataset as outside the 99.5% threshold (Fig. 3d; Table 3; Fig. S6) and consistent with in vitro and in vivo data, demonstrated linearity on the log-transformed scale (Fig. S6). Interestingly, this individual who did not conform to the model, had provocative clinical phenotype which included chronic spontaneous urticaria and angioedema as well as syncopal episodes with alcohol consumption that had prompted a bone marrow biopsy prior to genotyping. While bone marrow sections demonstrated left-shifted megaloblastoid erythroid hyperplasia, focal areas with increased pronormoblasts, and occasional dyserythropoietic forms, allele-specific PCR for *KIT* p.D816V was negative, no mast cell aggregates or aberrant markers (i.e., CD2, CD25) were identified, and no definitive diagnosis was made.

Finally to prospectively validate the modeled cut-off clinically, we measured BST levels in an additional 130 individuals with HαT caused by *TPSAB1* duplications in whom BST levels were unknown and no other clinical indication for bone marrow biopsy was present. In all 130 prospectively examined individuals, BST levels fell below the 99.5% cut-off (median 15.1 ng/mL, range 8.0-35.0 ng/mL).

### Application of tryptase genotyping and modeled BST levels improves biomarker utility

To further examine the real-world implications of applying this new model clinically, we analyzed all samples sent or referred to the NIH clinical center for tryptase genotyping from 2016-2021; 377 of these samples had not been used to construct the model. None of the participants had kidney disease or other clinical presentations consistent with genetic disorders known to be associated with elevated BST (13). Of these, 409 samples were identified with BST >11.4 ng/mL; 285 of these had HαT, 98 had MCD, 21 had both HαT and MCD, 2 individuals had hypereosinophilic disorders - one of whom also had HαT - and 4 individuals did not have a clear diagnosis at the time of genotyping. Thus, 98.8% (404/409) of individuals with BST >11.4 ng/mL could be accounted for by HαT and/or MCD (Fig. 3e). Moreover, 97.3% (255/262) of samples with BST levels between 8-11.4 ng/mL were associated with HαT and/or MCD, suggesting that even BST >8 ng/mL is uncommon among individuals without these conditions. Consistent with these data is the modeled 95% threshold among individuals without HαT or MCD of 7.99 ng/mL, indicating that less than 5% of the total population would be predicted to have BST >8 ng/mL.

The 5 individuals with BST >11.4 ng/mL who did not have HαT or evidence of MCD underwent paired peripheral blood exome and deep exome sequencing (300x) of granulocyte and mononuclear bone marrow aspirate fractions. In all five individuals, variants in genes commonly mutated in hematological malignancies (e.g., *TET2, RUNX1, CEBPA, MAP2K1, NOTCH2, KMT2C*) were identified (Table S6); in one individual this was associated with idiopathic hypereosinophilic syndrome. Together with those individuals meeting criteria for MCD, these data demonstrate that BST >11.4 ng/mL in the absence of HαT frequently identifies indolent clonal mast cell disease - 95% (98/103) of our referral cohort - or more rarely identifies patients with genetic variants suggestive of occult or evolving myeloid neoplasms.

Of note, 38.2% (55/144) samples with BST >20ng/mL were a result of HαT alone. Of those 55 participants, 54 were less than their respective threshold based on replication number. If tryptase genotyping was not considered, and instead the current minor criterion for SM of BST >20 ng/mL was used as a sole indication for bone marrow evaluation, 54 of these individuals could have been subjected to unnecessary invasive bone marrow examination. This is particularly relevant among individuals with two or more *TPSAB1* replications, where median BST levels were >20 ng/mL and can be >100 ng/mL, as seen in two related individuals with ten *TPSAB1* replications in cis (Fig. 2b, 3d; Table 3).

Finally, of the 21 individuals identified with both HαT and MCD (Table 4), 10 individuals had indolent SM (ISM), 8 of whom had BST levels above the 99.5% upper prediction limit dictated by *TPSAB1* replication number. Ten individuals had BST levels below the 99.5% upper prediction limit based upon *TPSAB1* replication number in our model; 6 with monoclonal mast cell activation syndrome (MMAS), 2 with ISM, 1 with diffuse cutaneous mastocytosis, and 1 with well-differentiated ISM. All these individuals had clinical implications for a bone marrow beyond the BST level and would have received a bone marrow regardless. Only one of these individuals - with the rare phenotype of well-differentiated ISM - would no longer have met clinical criteria for their diagnosis if the upper prediction limit was used as a minor criterion for the diagnosis of SM (rather than the current level of >20 ng/mL). Together, these data demonstrate that where tryptase genotyping is clinically available, these new reference ranges based upon *TPSAB1* replication number can be applied to the work-up of patients with elevated BST, establishing robust thresholds for which patients should undergo more extensive work-up including bone marrow aspiration and biopsy regardless of clinical presentation or symptomatology, and as an individualized minor criterion for the clinical diagnosis of SM.

**Table 4.**
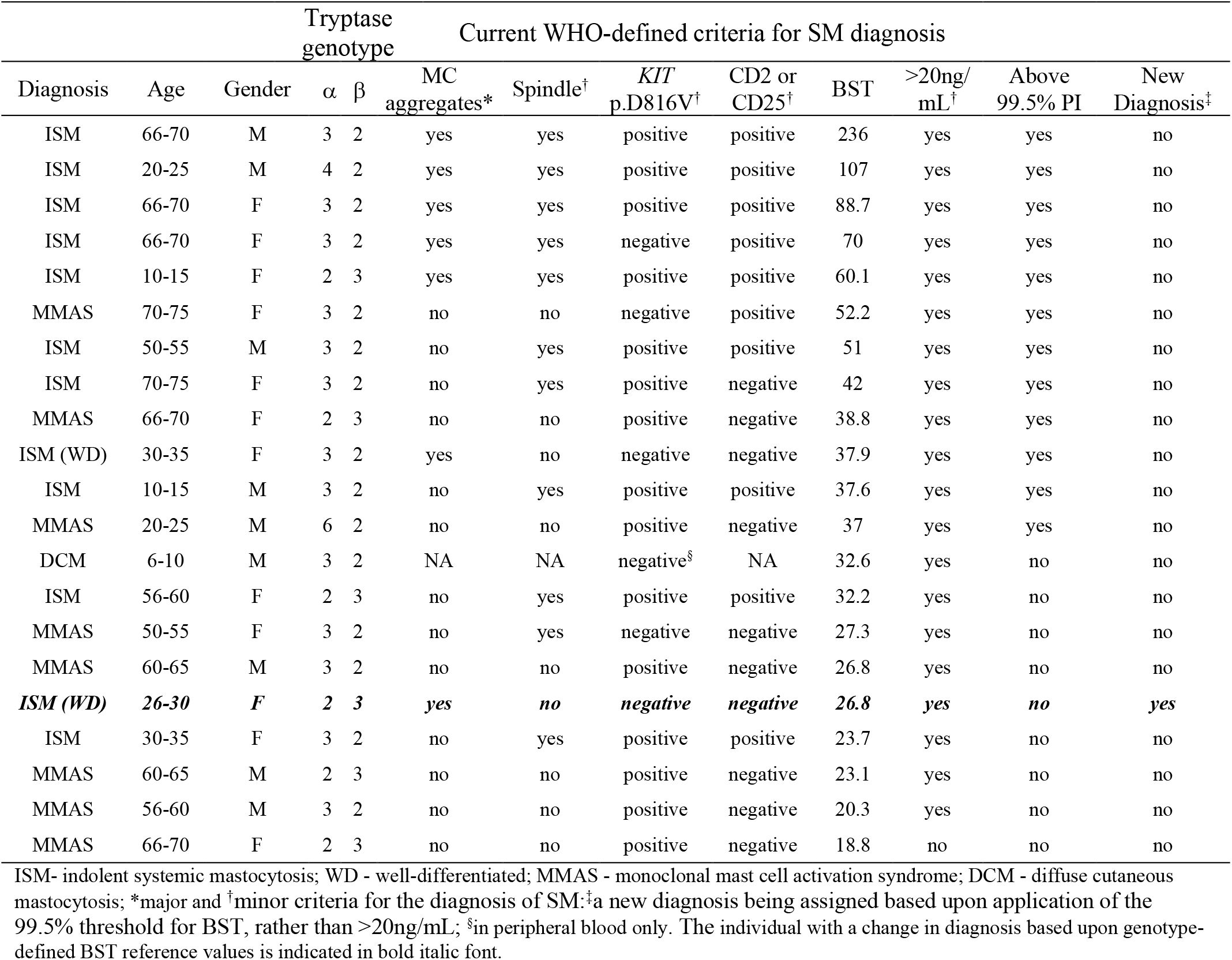
Individuals with MCD and HαT.

## DISCUSSION

Based upon our findings, HαT is a genetic trait best described as a naturally occurring over-expression model, whereby a conserved over-active protomer is co-inherited with α-tryptase-encoding *TPSAB1* gene replications driving basal over-expression of α^DUP^-tryptase that when translated, is indistinguishable from other α-tryptase proteins (i.e., α^WT^-tryptase). Using this knowledge and clinical data, we have generated a model that redefines clinically meaningful cut-offs for the upper limit of normal for serum tryptase levels based upon tryptase genotype. Moreover, we have demonstrated the clinical utility of using these individualized values in the evaluation of patients with elevated serum tryptase levels and in the diagnosis of systemic mastocytosis.

We acknowledge that MCD is not the only clonal myeloid neoplasm associated with elevated BST, rather it is the predominant one referred to our centers. Moreover, several studies have now demonstrated an increased prevalence of HaT among symptomatic individuals with SM (6-8) - likely due to potentiation of mast cell-mediated symptoms and reactions. Additional prospective studies will be helpful in validating the use of these new reference values in additional patients with HαT, MCD and other myeloid neoplasms. Severe renal dysfunction can also affect BST levels, as can other parasitic diseases which are rare in most Western countries (15). These are also not represented or evaluated in this study and should be considered when applying this model to patients. However, the model was generated based upon individuals without clinical signs or symptoms of these disorders. Thus, we expect these new reference values to remain robust in their predictive value, as has been suggested by a recent retrospective study of a regional health system where 93% (54/58) of individuals with BST >11.4 ng/mL had HαT, a myeloid neoplasm, or chronic kidney disease (22).

The data used to generate our model is largely from individuals with two or fewer replications. Thus, while available data for higher order copy number are consistent with our model, and indeed were incorporated into it, the generated cut-offs at high *TPSAB1* replication number (e.g., ≥3) are largely extrapolations. The a priori evidence demonstrated a linear relationship between BST and replication number providing support for this extrapolation. However, there is less certainty about the upper limits of BST level levels past two *TPSAB1* replications, and cut-offs for individuals with higher order copy number may require refinement as additional data become available.

To our knowledge, the application of genotypic information to determine clinical reference ranges in a personalized manner has not previously been used in laboratory medicine. However, many common clinical laboratory measures (e.g., immunoglobulin levels) are not normally distributed, and in some cases outliers may be similarly determined by heritable traits. Thus, we anticipate this kind of precision approach to clinical laboratory medicine will expand in the future as next generation sequencing becomes increasingly utilized in standard clinical practice.

## METHODS

### Study participants and samples

Patients, family members, and healthy volunteers provided informed consent on IRB-approved research protocols led by investigators with expertise in Allergy/Immunology at institutions that specialize in clonal and non-clonal mast cell disorders designed to study mastocytosis, or genetic diseases affecting the immune system at the NIH Clinical Center (NCT00852943, NCT01164241, NCT00044122, NCT00001756, NCT007197190), Antwerp University Hospital, Belgium (B300201525454), Verona University Hospital, Italy (protocol No. 39620), the University of Florida (IRB 201702274), the University of Mississippi Medical Center (IRB 2019-0082), or University Clinic Golnik, Slovenia (KME 150/09/13). All study participants had BST levels measured and tryptase genotyping performed (N=1,178). Among participants referred for evaluation or diagnosed with a mast cell-associated disorder (N=575), complete history and physical examinations were performed.

### Bone marrow biopsy and aspirate

Individuals presenting with signs or symptoms suggestive of a clonal mast cell disease (13) had additional clinical work-up that included bone marrow biopsy and aspirate. Immunohistochemistry of bone marrow sections was performed for enumeration and characterization of mast cells (KIT and tryptase), and evaluation of CD2 and CD25 expression in aspirate and/or tissue section. Allele-specific PCR for *KIT* p.D816V in peripheral blood and bone marrow was also performed in these patients. In the rare case that an individual declined bone marrow biopsy, peripheral blood *KIT* p.D816V was performed by allele-specific or ddPCR. If the result was positive, these individuals were considered to have clonal mast cell disease and included. Whereas, if *KIT* p.D816V screening of peripheral blood was negative in an individual with an incomplete work-up for a suspected myeloid neoplasm, such individuals were excluded from the study.

### Total basal serum tryptase quantification

Total basal serum tryptase (BST) levels were measured using the commercially available ImmunCAP assay (Thermo Fisher Scientific, Phadia AB, Uppsala, Sweden) or ELISA as described (16), performed in CLIA-certified laboratories (Mayo Clinic, Rochester, NY and Virginia Commonwealth University, Richmond, VA).

### Bacterial artificial chromosome (BAC) library generation

Genomic DNA was isolated from PBMC of an individual homozygous for ααβ tryptase alleles. HMW DNA fragments were then partially digested with the restriction enzyme BamHI and size selected prior to ligation of fragments into the pCC1BAC vector (Epicentre Biotechnologies, Madison, WI) and transformation of DH10B *E. coli* cells (17, 18). The clones were robotically chosen and replicated. Replicated clone copies were used as a source plate for constructing nylon filters. The filter hybridization was carried out using the DIG method for filter hybridization as described (19) (Amplicon Express, Pullman, WA). Verification of putative clones was carried out by conventional and digital droplet PCR (ddPCR).

### BAC sequencing and assembly

Genomic DNA from selected clones was isolated (20) and sequenced using Single-molecule real-time sequencing (SMRT) sequencing technology (PacBio, Menlo Park, CA) at DNA Link (South Korea). De novo assembly was accomplished using HGAP2 and the Canu assembler for PacBio on SMRT Analysis platform version 2.2.0 and the SMRTPortal: SMRTAnalysis build 133377, Daemon version v2.2.0 build 132105, SMRTpipe version v2.2.0 build 132739, SMRT Portal version v2.2.0 build 133335, SMRT View version v2.2.0 build 132578.

### Bioinformatic analyses

#### Paired-read tryptase haplotype analysis

Genome sequence reads mapping to the tryptase locus were re-aligned to a 142-bp region (Table S1). Detailed alignments for each read were parsed to extract the sequence at each of the five specified variable positions if covered by the read. Haplotype assignments were determined based on the collection of positional sequence assignments determined for each read pair. Generated haplotypes were then correlated to tryptase genotype and linkage to inherited alleles was confirmed when pedigrees were available.

#### RNA-seq dataset analyses

Using a 39-bp consensus sequence (Table S2), gene expression datasets (see Accession codes for list of datasets) were reanalyzed using NCBI Magic-BLAST (https://ncbi.github.io/magicblast/), in order to map reads to 3 gene isoforms: α^DUP^-, α^WT^-, and β-tryptase. Samples were designated as tryptase-positive, tryptase-negative, α^DUP^-, α^WT^-, and β-tryptase only based on the criteria of having qualified reads that mapped exactly without any mismatches to the interval between 25 bp-45 bp from 5’ end of the transcripts (see Supplemental Methods).

### Promoter amplification and cloning

Promoter amplicons were generated from genomic DNA from individuals with increased *TPSAB1* copy number using primers designed on conserved sequences present in all identified alpha- and beta-tryptase sequences (FWD – GGGCAAGTCCACAGGGAGCT; REV – CTGGGGAGCAAGGAGGAGCA) in order to amplify all sequences between the ATG start site and a conserved region ∼1-2 kb 5’ of the variably expanded repeat region (Fig. 1). Amplification was confirmed by gel electrophoresis and clones of products were generated using the TOPO^®^ Cloning Kit (Thermo, Waltham, MA) and transformed into One Shot® TOP10 Chemically Competent E. coli (Thermo). Single colonies were selected, confirmed to contain single intact clones by PCR and gel electrophoresis, and Sanger sequenced using the FWD and REV, as well as two additional internal primers (TGCAGGTGCAACCCCAGGA and TCCTGGGGTTGCACCTGCA).

For α^DUP^- and α^WT^-tryptase-specific promoter cloning, the reverse primer (GACGATACCCGCTTGCTGCAG) was located across alpha-tryptase-specific exonic sequence used for the isoform-specific genotyping assay, and for α^WT^-tryptase the universal FWD primer was used and for α^DUP^-tryptase a different sequence normally only seen linked to beta-tryptase was used (GGGCAAGTCCACAGGGAGCT). Resulting amplicon size was confirmed by gel electrophoresis (Fig. S2).

### Reporter assay

Sequence-validated clones corresponding to α^DUP^- and α^WT^-tryptase promoters identified in the BAC assembly were subcloned into the reporter plasmid pDD-AmCyan1 (Takara Bio USA, Inc., Mountain View, CA) and Sanger sequence verified. MonoMac-6 cells were transfected with the reporter plasmid pDD-AmCyan1 containing α^DUP^- or α^WT^-tryptase promoter clones by electroporation using Cell Line Nucleofector™ Kit V (Lonza, Basel, Switzerland) using the setting U-005 according to manufacturer instructions. Cells were cultured in standard media in the presence of Shield1 (Takara Bio) according to manufacturer instructions and basal fluorescence was measured at indicated time points and recorded using an LSR Fortessa (BD Biosciences) and analyzed using FlowJo (Treestar, Ashland, OR).

### Basophil isolation

Following isolation of PBMC via density centrifugation, basophils were negatively selected using the MACS^®^ Diamond Basophil Isolation Kit (Miltenyi Biotec, San Diego, CA).

### Cell culture

#### Primary mast cells

Peripheral blood mononuclear cells (PBMCs) were isolated using density centrifugation and CD34^+^ cells were positively selected using the ferromagnetic bead-based MACS^®^ system (Miltenyi) and the number of cells was quantified. Mast cells were then differentiated under the established conditions described (21).

#### Cell lines

MonoMac-6 cells were cultured in RPMI 1640 + 10% FBS + 2 mM L-glutamine + non-essential amino acids + 1 mM sodium pyruvate + 10 µg/ml human insulin).

### Droplet digital PCR (ddPCR)

#### Tryptase genotyping

Alpha- and beta-tryptase sequences at *TPSAB1* and *TPSB2* were genotyped as described (5) using droplet digital PCR (ddPCR). Briefly, genomic DNA was extracted from PBMCs, cell lines, or obtained from the HapMap biorepository and restriction endonuclease treated. Custom primer/probe sets specific for alpha- and beta-tryptase were then employed using the PrimePCR ddPCR copy number reference *AP3B1*, according to the manufacturer’s specifications (Bio-Rad; Hercules, CA).

#### Gene expression

Total RNAs were extracted using Trizol reagent (Thermo) and the RNEasy Mini Kit (Qiagen) from basophils, primary cultured mast cells, or cell lines under stated conditions and reverse-transcribed using SuperScript III First-Strand Synthesis System (Thermo). ddPCR was performed on a QX200 system (Bio-Rad) according to the manufacturer’s instructions using custom primer/probe sets for tryptase isoforms as indicated (Table S3) to quantify gene expression, normalizing to *HPRT1* and *TBP* expression quantified using commercially available ddPCR primer/probe sets (Bio-Rad).

#### Promoter detection assay

A custom primer/probe set was designed to amplify the proximal promoter linked to alpha-tryptase (Fig. S4). The forward primer corresponds to a conserved sequence present in all identified α- and β-tryptases. The reverse primer hybridizes to sequences only present in alpha-tryptases. The probes compete for hybridization with the C>T variant that defines α^DUP^-, and α^WT^-tryptase-specific sequences, respectively. To successfully amplify the large 71% GC-rich amplicon, the reaction was run with 10% 1M Betaine (Sigma, Saint Louis, MO) under the following conditions: 95 °C 10 min, 96 °C 30 s, 66 °C 1 min (Ramp 1.5 °C), 50 cycles, 96°C 10 min.

### Exome sequencing and bioinformatic analysis for clonal variants

Bone marrow aspirates were fractionated using density centrifugation into granulocyte and mononuclear fractions using a double gradient 1.077 g/ml (Histopaque-1077) and 1.119 g/ml (Histopaque-1119) (Sigma, St. Louis, MO). Genomic DNA was extracted and libraries were prepared with the Twist Biosciences Comprehensive Exome capture kit. Deep (>300x) exome sequencing was subsequently performed using the Illumina HiSeq 2500, on both the granulocyte and mononuclear bone marrow fractions. Paired peripheral blood samples from the same individuals were also exome sequenced consistent with clinically recommended standards (22).

Raw fastq files were trimmed for quality and adapter contamination using Trimmomatic v0.39 (23) and mapped to the human hg38 reference genome using BWA-MEM v0.7.17 (http://bio-bwa.sourceforge.net/). PCR duplicates were marked using Samlaster v0.1.2.5 (24), and GATK v4.1.9.0 was used to perform base recalibration. For the peripheral blood samples, germline variation was called using GATK v4.1.9.0 and following the recommended Best Practices (https://gatk.broadinstitute.org/hc/en-us/articles/360035535932-Germline-short-variant-discovery-SNPs-Indels-). For granulocyte and mononuclear fractions, somatic variants were called using 2 approaches. First, paired somatic calling with the PBMC sample treated as germline was performed using 3 somatic detection tools: 1) MuTect2 from GATK v4.1.9.0 and following the recommended Best Practices (https://gatk.broadinstitute.org/hc/en-us/articles/360035531132--How-to-Call-somatic-mutations-using-GATK4-Mutect2), 2) Strelka v2.9.0 (25), and 3) mutect v1.1.7 (26). In addition to paired calling, we also performed tumor-only calling with a combination of MuTect2 GATK v4.1.9.0, mutect v1.1.7, and Vardict v1.7.0 (27). The latter approach of performing somatic variant detection without including the paired PBMC sample was done to account for the fact that somatic variants in the bone marrow granulocytes and bone marrow mononuclear cells could also be present at low fractions in the PBMC sample and therefore get excluded as contaminating germline variants in the paired calling. For both approaches, variants were then merged across all callers and annotated with VEP v104 (28) for downstream analysis. Reported variants were prioritized based upon commonly implicated genes in the development of myeloid neoplasms.

### Statistical modeling and analyses

To predict BST levels based upon tryptase genotypes - specifically determined by *TPSAB1* replication number (including those without replications) - a log transformation was applied to the BST levels because the values were not normally distributed. Additionally, we log-transformed the replication number, because transforming BST back to the normal scale resulted in a model fit that was approximately linear, with a near constant increase in BST for each increase in replication number; this supported results from early clinical data and data generated from in silico and in vitro experiments. We also added 0.5 to replication number prior to log transformation in order to include individuals without *TPSAB1* replications in the model, thus allowing us to also determine a cutoff for individuals without HαT. Thus, we used a linear regression model to predict log(BST) from log(replication number plus 0.5), and from that, created an upper one-sided 99.5% prediction interval. The 99.5% threshold was chosen as it provides high specificity where very few individuals will be identified as false positives and correlated well with the predicted prevalence of elevated BST in the absence of HαT based upon available population data (10). The model was developed using R version 3.6.3.

For clinical and experimental data, Mann-Whitney, Kruskal-Wallis, and paired and unpaired two-tailed t-tests, were used where appropriate to test significance of differences, prevalence, or deviation using Prism (GraphPad).

### Basal Serum Tryptase (BST) calculator code

We developed an online calculator the **<u>B</u>**asal **<u>S</u>**erum **<u>T</u>**ryptase **<u>C</u>**linical cut-off **<u>A</u>**ssigned by **<u>L</u>**ocus **<u>C</u>**opy number of **<u>U</u>**TR-**<u>L</u>**inked element and **<u>A</u>**ssociated **<u>*T*</u>***PSAB1* **<u>E</u>**ncoded **<u>R</u>**eplication (BST CALCULATER) with Shiny R framework: https://bst-calculater.niaid.nih.gov/. The code is available at: https://github.com/niaid/BST-calculater

### URLs

Burrows-Wheeler Aligner **http://bio-bwa.sourceforge.net/**

Picard, **http://broadinstitute.github.io/picard/**

Plink, **http://pngu.mgh.harvard.edu/purcell/plink/**

Genome Analysis Toolkit (GATK), **http://www.broadinstitute.org/gatk/**

SAMtools, **http://www.htslib.org/**

### Accession Codes

Sequencing data were obtained from the National Center for Biotechnology Information Sequence Read Archive for the following BioProjects: hereditary alpha-tryptasemia: **PRJNA342304**; individuals not selected by tryptase genotype: **PRJNA208369 PRJNA219425 PRJNA232669 PRJNA252605 PRJNA253059 PRJNA254943 PRJNA257389 PRJNA258216 PRJNA261011 PRJNA261251 PRJNA263242 PRJNA266512 PRJNA266572 PRJNA270371 PRJNA271942 PRJNA274028 PRJNA274360 PRJNA275801 PRJNA279249 PRJNA280990 PRJNA283839 PRJNA289905 PRJNA291619 PRJNA292690 PRJNA293555 PRJNA296379 PRJNA301173 PRJNA301364 PRJNA310988 PRJNA315611 PRJNA317535 PRJNA318253 PRJNA319220 PRJNA326113 PRJNA327986 PRJNA330840 PRJNA340161 PRJNA342177 PRJNA343985 PRJNA354367 PRJNA358081 PRJNA369563 PRJNA369684 PRJNA373887 PRJNA376200 PRJNA377555 PRJNA378385 PRJNA384963 PRJNA388978 PRJNA389466 PRJNA392116 PRJNA395367 PRJNA395589 PRJNA399103 PRJNA400331 PRJNA407731 PRJNA415746 PRJNA428940 PRJNA214592 PRJNA229548 PRJNA278364 PRJNA278767 PRJNA263397**

### Referenced GenBank Accession Codes

GRCh37/hg19 NCBI assembly: **GCF_000001405.13**; tryptase locus assemblies: **AC226137.3 AC120498.2 CTD-2503P16 CHM13 AL031704.24 AE006466.1 CHM1 AC240106.3 AC238650.2 AC213746.1 GM24385 NA12878 AF098328.1**

## Data Availability

All data produced in the present study are available upon reasonable request to the authors.

## Author Contributions

J.J.L. designed the study. J.J.L., S.L., C.N., J.B., T.D., M.C.C., H.D.K., J.S., K.M.H., E.F., P.K., J.D.M., V.S., D.E., P.B., R.Z., P.K., and D.D.M, recruited and enrolled study participants. J.J.L., I.T., J.H., J.H., J.M., C.S.H., B.B., M.P., N.N.T., R.E.H., S.A.M., G.H.C., and J.L. performed and/or supervised the bioinformatics analyses. J.J.L. designed and J.C., J.K., and Y.L. performed ddPCR assays. I.M. performed hematopathologic analyses. J.J.L., J.C., J.K., D.D.M., Y.B., Y.Y., Y.H.P., Y.L., M.P.O., and A.P.H. designed, supervised, and /or performed the functional and genetic studies and sequencing, respectively. A.M. and E.H.B. developed the prediction interval model. J.J.L. prepared the draft manuscript. All authors contributed to discussion of the results and to manuscript preparation.

## Acknowledgements

The investigators thank the patients, their families, and healthy volunteers who contributed to this research. We also thank Adam Phillippy for sharing his expertise on complex genetic structural rearrangement.

