## Supplemental Materials for "Genetically determining individualized clinical reference ranges for the biomarker tryptase can limit unnecessary procedures and unmask myeloid neoplasms"

### SUPPLEMENTAL METHODS

### Tryptase isoform re-alignment script

```
import pysam
import glob
import pandas as pd
import argparse

parser = argparse.ArgumentParser(description='Create a CSV file for further processing')

parser.add_argument('--bam_folder', type=str, nargs=1, required=True,
                    help='The directory containing the bam files.')
parser.add_argument('--source', type=str, nargs=1, required=True,
                    help='The source of the bam files e.g. AML')
parser.add_argument('--roi_start', type=int, nargs=1, required=True,
                    help='The 0 based start position of ROI')
parser.add_argument('--roi_end', type=int, nargs=1, required=True,
                    help='The 0 based end position of ROI')
parser.add_argument('--output_name', type=str, nargs=1, required=True,
                    help='The name of the output file')
args = parser.parse_args()

print (args)

def get_files(directory_path):
    """
    Return a list containing all the files matching a pattern in \
    the given directory

    """

    print ('{directory_path}/*.sorted.bam'.format(
                    directory_path=directory_path))
    file_list  = glob.glob('{directory_path}/*.sorted.bam'.format(
                    directory_path=directory_path))

    return file_list

def get_read_count(df):
    """
    Get the read count in a specific bam file.

    """

    return int(pysam.view("-c", df['file_location']))

def get_transcript_count(df, transcript_name):

    """
    Return the count of the transcript i.e how many of each of \
    the three transcripts are in there.
    Note - Does not look at the quality of the alignment.
    """

    sam_file_location = df['file_location']

    samfile = pysam.AlignmentFile(sam_file_location, "rb")

    count = 0

    for read in samfile:

        if read is not None and read.reference_name == transcript_name:

            count = count + 1

    return count

def get_zero_edit_distance_count(df, transcript_name):

    """
    For a goven transcript e.g. Alpha_GEX_64k_HEX get how many of \
    the matches have an NM tag of zero
    i.e. the match was exact.

    """

    sam_file_location = df['file_location']

    samfile = pysam.AlignmentFile(sam_file_location, "rb")

    count = 0

    for read in samfile:

        if read is not None and read.reference_name == transcript_name:

            edit_distance = read.get_tag('NM')

            if edit_distance == 0:

                count = count + 1

    return count

def get_transcript_read_count_filtered(df, transcript_name, start, end):

    """
    Count hits which cover the bit of the reference we are interested in.

    start = 0 based position in reference the alignment must start on or before.
    end = 0 based position that the alignment must end on or before.

    """

    sam_file_location = df['file_location']

    samfile = pysam.AlignmentFile(sam_file_location, "rb")

    iter = samfile.fetch(transcript_name, start, end)

    count =0

    for read in iter:

        if read.reference_start <=start and read.reference_end >= end:

            count = count +1

    return count

def get_transcript_read_count_filtered_exact(df, transcript_name, start, end):

    """
    Count hits which cover the bit of the reference we are interested in \
    and which are exact.

    That is - do they cross position 0 - 44 of the transcript \
    (given by transcipt_name) and \
    have an edit distance from the reference of 0.

    start = 0 based position in reference the alignment must start on or before.
    end = 0 based position that the alignment must end on or before.

    """

    sam_file_location = df['file_location']

    samfile = pysam.AlignmentFile(sam_file_location, "rb")

    iter = samfile.fetch(transcript_name, start, end)

    count =0

    for read in iter:

        if read.reference_start <=start and read.reference_end >=end:

            edit_distance = read.get_tag('NM')

            if edit_distance == 0:

                count = count + 1

    return count

results = get_files(args.bam_folder[0])
file_names = [x.split('/')[len(x.split('/'))-1] for x in results]

#Create Dataframe
df = pd.DataFrame(index=file_names)

df['file_location'] = results
df['source'] = args.source[0]

#Add total-count column
df['alignment_count'] = df.apply(get_read_count, axis=1)
print ('Added total alignment count')

#Count the number of reads aligned to each reference transcript
df['alpha_wt_count'] = df.apply(get_transcript_count, axis=1, args=['Alpha_GEX_64k_HEX'])
df['alpha_dup_count'] = df.apply(get_transcript_count, axis=1, args=['Alpha_GEX_79k_dup_FAM'])
df['beta_count'] = df.apply(get_transcript_count, axis=1, args=['BETA_new_GEX_FAM'])
print ('Added alignment count for each transcript')

#Number of exact read alignments for each reference transcript
df['alpha_wt_zero_edit_count'] = df.apply(get_zero_edit_distance_count, axis=1, args=['Alpha_GEX_64k_HEX'])
df['alpha_dup_zero_edit_count'] = df.apply(get_zero_edit_distance_count, axis=1, args=['Alpha_GEX_79k_dup_FAM'])
df['beta_zero_edit_count'] = df.apply(get_zero_edit_distance_count, axis=1, args=['BETA_new_GEX_FAM'])
print ('Added exact alignment count for each transcript')

# Number of alignments that span our area of interest

start = args.roi_start[0]
end = args.roi_end[0]

df['alpha_read_covers_snps_count'] = df.apply(get_transcript_read_count_filtered,
                                              axis=1,
                                              args=['Alpha_GEX_64k_HEX', start,end])
df['alpha_dup_read_covers_snps_count'] = df.apply(get_transcript_read_count_filtered,
                                              axis=1,
                                              args=['Alpha_GEX_79k_dup_FAM', start,end])
df['beta_read_covers_snps_count'] = df.apply(get_transcript_read_count_filtered,
                                              axis=1,
                                              args=['BETA_new_GEX_FAM', start,end])
print ('Added alignment count for each transcript within ROI')

# Number of alignments that span our area of interest and are exact
df['alpha_read_covers_snps_count_exact'] = df.apply(get_transcript_read_count_filtered_exact,
                                              axis=1,
                                              args=['Alpha_GEX_64k_HEX', start,end])

df['alpha_dup_read_covers_snps_count_exact'] = df.apply(get_transcript_read_count_filtered_exact,
                                              axis=1,
                                              args=['Alpha_GEX_79k_dup_FAM', start,end])

df['beta_read_covers_snps_count_exact'] = df.apply(get_transcript_read_count_filtered_exact,
                                              axis=1,
                                              args=['BETA_new_GEX_FAM', start,end])

print ('Added exact alignment count for each transcript within ROI')

df.to_csv(args.output_name[0])
print ('Created CSV')
```

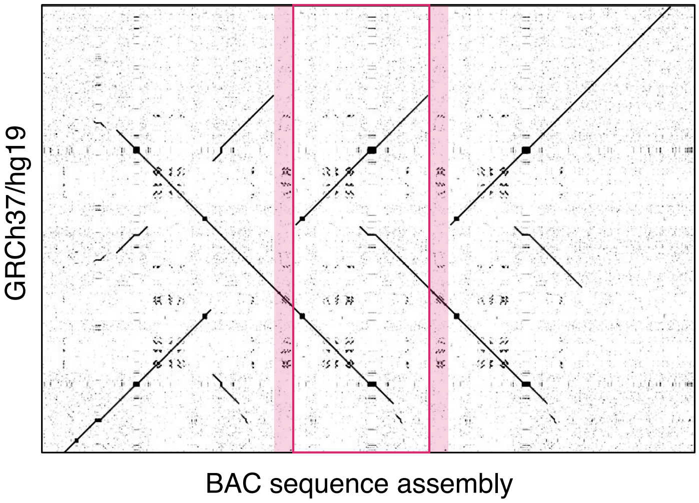

**Figure S1. Alignment of the tryptase locus in hereditary alpha tryptasemia.** Dot-plot of the assembled tryptase locus from an individual with a *TPSAB1* duplication (BAC clone assembly, X-axis) with the human reference sequence GRCh37/hg19 chr16:1,270,000-1,315,000 (Y-axis).

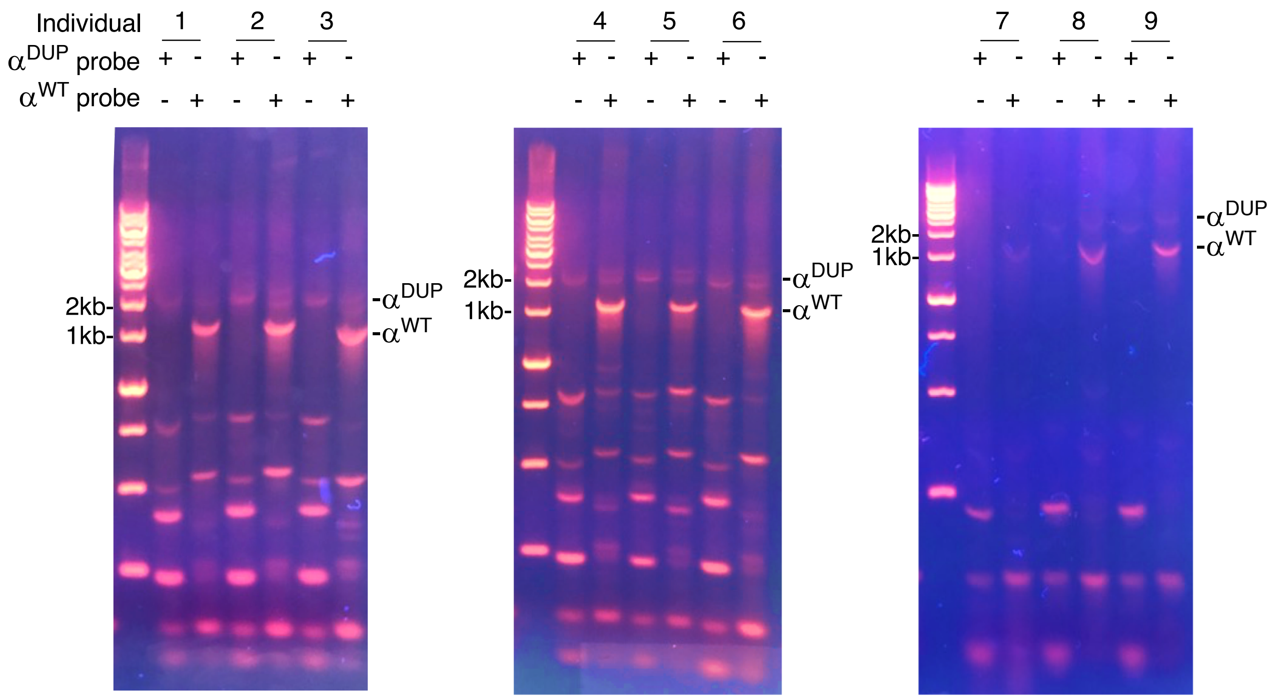

**Figure S2. Gel electrophoresis of alpha-tryptase linked promoters.** Genomic DNA from nine individuals with hereditary alpha tryptasemia was amplified with duplicated ($\alpha^{DUP}$) and wild-type alpha-tryptase ($\alpha^{WT}$) specific forward primers paired with an alpha-tryptase specific reverse primer**.** The largest molecular weight bands of approximately 1 and 2 kilobases (kB) respectively correspond to the full-length proximal coding sequences and linked promoters.

```
CLUSTAL O(1.2.4) multiple sequence alignment

aDUP_CACCT_BAC          GGGGCCGGGGGTGAGACCATGGGGAGCTGGGCTGGGGCTGGGGGTGAGACCATGGGGAGC     60
aDUP_CACCT_CLONE8       GGGGCCGGGGGTGAGACCATGGGGAGCTGGGCTGGGGCTGGGGGTGAGACCATGGGGAGC     60
aWT_GGTTC_BAC           GGGGCCGGGGATGAGACCATGGGGAGCTGGG-----------------------------     0
aWT_GGTTC_CLONE5        GGGGCCGGGGATGAGACCATGGGGAGCTGGG-----------------------------     0
                        **********.********************

aDUP_CACCT_BAC          TGGGGCCGGGGGCTGGGACTAGTCCATGGGGAGCTGGGCTGGGGCTGGGGGTGAGACCAT     120
aDUP_CACCT_CLONE8       TGGGGCCGGGGGCTGGGACTAGTCCATGGGGAGCTGGGCTGGGGCTGGGGGTGAGACCAT     120
aWT_GGTTC_BAC           ------------------------------------------------------------     0
aWT_GGTTC_CLONE5        ------------------------------------------------------------     0

aDUP_CACCT_BAC          GGGGAGCTGGGCTGGGGCTGGGACTAGTCCATGGGGAGCTGGGCTGGAGCTGGGGGTGAG     180
aDUP_CACCT_CLONE8       GGGGAGCTGGGCTGGGGCTGGGACTAGTCCATGGGGAGCTGGGCTGGAGCTGGGGGTGAG     180
aWT_GGTTC_BAC           ------------------------------------------------------------     0
aWT_GGTTC_CLONE5        ------------------------------------------------------------     0

aDUP_CACCT_BAC          ACCATGGGGAGCTGGGGCCGGGGGCTGGGACTAGTCCATGGGGAGCTGGGCTGGGGCTGG     240
aDUP_CACCT_CLONE8       ACCATGGGGAGCTGGGGCCGGGGGCTGGGACTAGTCCATGGGGAGCTGGGCTGGGGCTGG     240
aWT_GGTTC_BAC           ------------------------------------------------------------     0
aWT_GGTTC_CLONE5        ------------------------------------------------------------     0

aDUP_CACCT_BAC          GGGTGAGACCATGGGGAGCTGGGCTGGGGCTGGGACTAGTCCATGGGGAGCTGGGGGTGA     300
aDUP_CACCT_CLONE8       GGGTGAGACCATGGGGAGCTGGGCTGGGGCTGGGACTAGTCCATGGGGAGCTGGGGGTGA     300
aWT_GGTTC_BAC           ------------------------------------------------------------     0
aWT_GGTTC_CLONE5        ------------------------------------------------------------     0

aDUP_CACCT_BAC          GACCATGGGGAGCTGGGGCCGGGGGCTGCGACTAGTCCATGGGGAGCTGGGCTGGGGCTG     360
aDUP_CACCT_CLONE8       GACCATGGGGAGCTGGGGCCGGGGGCTGCGACTAGTCCATGGGGAGCTGGGCTGGGGCTG     360
aWT_GGTTC_BAC           ------------------------------------------------------------     0
aWT_GGTTC_CLONE5        ------------------------------------------------------------     0

aDUP_CACCT_BAC          GGGGTGAGTCCATGGGGAGCTGGGCTGGGGCTGGGGGTGAGACCATGGGGAGCTGGGCTG     420
aDUP_CACCT_CLONE8       GGGGTGAGTCCATGGGGAGCTGGGCTGGGGCTGGGGGTGAGACCATGGGGAGCTGGGCTG     420
aWT_GGTTC_BAC           ------------------------------------------------------------     0
aWT_GGTTC_CLONE5        ------------------------------------------------------------     0

aDUP_CACCT_BAC          GGGCTGGGGGTGAGACCATGGGGAGCTGGGGCCGGGGGATGGGACTAGTCCATGGGGAGC     480
aDUP_CACCT_CLONE8       GGGCTGGGGGTGAGACCATGGGGAGCTGGGGCCGGGGGATGGGACTAGTCCATGGGGAGC     480
aWT_GGTTC_BAC           ------------------------------------------------------------     0
aWT_GGTTC_CLONE5        ------------------------------------------------------------     0

aDUP_CACCT_BAC          TGGGCTGGGGCTGGGGGTGAGACCATGGGGAGCTGGGCTGGGGCTGGGACTAGTCCATGG     540
aDUP_CACCT_CLONE8       TGGGCTGGGGCTGGGGGTGAGACCATGGGGAGCTGGG-----------------------     517
aWT_GGTTC_BAC           ------------------------------------------------------------     31
aWT_GGTTC_CLONE5        ------------------------------------------------------------     31

aDUP_CACCT_BAC          GGAGCTGGGCTGGAGCTGGGGGTGAGACCATGGGGAGCTGGGGCCGGGGGCTGCGACTAG     600
aDUP_CACCT_CLONE8       ------------------------------------------------------------     517
aWT_GGTTC_BAC           ------------------------------------------G-----------------     32
aWT_GGTTC_CLONE5        ------------------------------------------G-----------------     32

aDUP_CACCT_BAC          TCCATGGGGAGCTGGGCTGGGGCTGGGGGTGAGTCCATGGGGAGCTGGGCTGGGGCTGGG     660
aDUP_CACCT_CLONE8       -------------------------------------------------CTGGGGCTGGG     528
aWT_GGTTC_BAC           -------------------------------------------------CTGGGGCT-GG     42
aWT_GGTTC_CLONE5        -------------------------------------------------CTGGGGCT-GG     42
                                                                         ******** **

aDUP_CACCT_BAC          GGTGAGTCCATGGGGAGCTGGGGCTGGGGCTGGGACTAGTCCATGGGCAGCTGGGCTGGG     720
aDUP_CACCT_CLONE8       GGTGAGTCCATGGGGAGCTG----------------------------------------     548
aWT_GGTTC_BAC           GACTAGTCCATGGGGAGCTGG---------------------------------------     63
aWT_GGTTC_CLONE5        GACTAGTCCATGGGGAGCTGG---------------------------------------     63
                        *.  ****************

aDUP_CACCT_BAC          GCTGGGGGTGAGTCCATGGGGAGCTGGGCTGGGGCTGGGGGTGAGTCCATGGGGAGCTGG     780
aDUP_CACCT_CLONE8       -----------------------------------------------------------G     549
aWT_GGTTC_BAC           -----------------------------------------------------------G     64
aWT_GGTTC_CLONE5        -----------------------------------------------------------G     64
                                                                                   *

aDUP_CACCT_BAC          GCTGGGGTTGGGGGTGAGTCCATGGGGAGCTGGGCTGGGTCTCCTGGGGTTGCACCTGCA     840
aDUP_CACCT_CLONE8       GCTGGGGTTGGGGGTGAGTCCATGGGGAGCTGGGCTGGGTCTCCTGGGGTTGCACCTGCA     609
aWT_GGTTC_BAC           GCTGAAGTTGGGGGTGAGTCCATGGGGAGCTGGGCTGGGCCTCCTGGGGTTGCACCTGCA     124
aWT_GGTTC_CLONE5        GCTGAAGTTGGGGGTGAGTCCATGGGGAGCTGGGCTGGGCCTCCTGGGGTTGCACCTGCA     124
                        ****..********************************* ********************

aDUP_CACCT_BAC          CTCCTGTCTGCCCTTCCCTCTGCCGATGAAGCTCAGATCCCATGATAAGGAGGCACCTGC     900
aDUP_CACCT_CLONE8       CTCCTGTCTGCCCTTCCCTCTGCCGATGAAGCTCAGATCCCATGATAAGGAGGCACCTGC     669
aWT_GGTTC_BAC           CTCCTCTCTGCTCTTCCCTCTGCGTATGAAGCTCAGATCCCATGATAAGGAGGCACCTGC     184
aWT_GGTTC_CLONE5        CTCCTCTCTGCTCTTCCCTCTGCGTATGAAGCTCAGATCCCATGATAAGGAGGCACCTGC     184
                        ***** ***** ***********  ***********************************

aDUP_CACCT_BAC          AGACCAGGGGTCCTGCACGGACAGCCCCAGAGGTGGACATTGAGGACTCGTAGGAGGACT     960
aDUP_CACCT_CLONE8       AGACCAGGGGTCCTGCACGGACAGCCCCAGAGGTGGACATTGAGGACTCGTAGGAGGACT     729
aWT_GGTTC_BAC           AGACCAGGGGACCTGCACGGACAGCCCCAGAGGTGGACATTGAGGACTCGTAGGAGGACT     244
aWT_GGTTC_CLONE5        AGACCAGGGGACCTGCACGGACAGCCCCAGAGGTGGACATTGAGGACTCGTAGGAGGACT     244
                        **********:*************************************************

aDUP_CACCT_BAC          TGGGTCTCATACGGCGGGTGGGGAGCAGGGCCCCTTCCTGGCTGAGGACACTTGGTGCTG     1020
aDUP_CACCT_CLONE8       TGGGTCTCATACGGCGGGTGGGGAGCAGGGCCCCTTCCTGGCTGAGGACACTTGGTGCTG     789
aWT_GGTTC_BAC           TGGGTCTCATACGGCGGGTGGGGAGCAGGGCCCCTTCCTGGCTGAGGACACTTGGTGCTG     304
aWT_GGTTC_CLONE5        TGGGTCTCATACGGCGGGTGGGGAGCAGGGCCCCTTCCTGGCTGAGGACACTTGGTGCTG     304
                        ************************************************************

aDUP_CACCT_BAC          TCCCCTCTCAAGGCTGTTTCCCCATCTGACAAAGGGGTCTCATGTGAGCCTCCCACCAAG     1080
aDUP_CACCT_CLONE8       TCCCCTCTCAAGGCTGTTTCCCCATCTGACAAAGGGGTCTCATGTGAGCCTCCCACCAAG     849
aWT_GGTTC_BAC           TCCCCTCTCAAGGCTGTTTCCCCATCTGACAAAGGGGTCTCATGTGAGCCCCCAACCAAG     364
aWT_GGTTC_CLONE5        TCCCCTCTCAAGGCTGTTTCCCCATCTGACAAAGGGGTCTCATGTGAGCCCCCAACCAAG     364
                        ************************************************** **.******
```

```
aDUP_CACCT_BAC        TGAGTCGAGGAGGGCTGGCGCCACCCCCGTGGATTCGGAGTCCGTAAGAGGGGTGTCACC    1140
aDUP_CACCT_CLONE8     TGAGTCGAGGAGGGCTGGCGCCACCCCCGTGGATTCGGAGTCCGTAAGAGGGGTGTCACC    909
aWT_GGTTC_BAC         TGAGTCGAGGAGGGCTGGCCCCACCCCCGTGGATTTGGAGTCCGTAGGAGGGGTGTCACC    424
aWT_GGTTC_CLONE5      TGAGTCGAGGAGGGCTGGCCCCACCCCCGTGGATTTGGAGTCCGTAGGAGGGGTGTCACC    424
                      ******************* *************** **********.*************

aDUP_CACCT_BAC        CGTCATGTCCCCACCCCGTGGGCACCTTCCCGTCTCTTGGAGGGTGGCCCATGGACATGA    1200
aDUP_CACCT_CLONE8     CGTCATGTCCCCACCCCGTGGGCACCTTCCCGTCTCTTGGAGGGTGGCCCATGGACATGA    969
aWT_GGTTC_BAC         CGTCATGTCCCCACCCCGTGGGCACCTTCCCGTCTCTTGGAGCGTGGCCCATGGACATGA    484
aWT_GGTTC_CLONE5      CGTCATGTCCCCACCCCGTGGGCACCTTCCCGTCTCTTGGAGCGTGGCCCATGGACATGA    484
                      ****************************************** *****************

aDUP_CACCT_BAC        GTTCCTCACCCCGTGTCCCTCTTGGGGAAACAGGTTTCAGGAGCGATGGGTCTTGTAGCC    1260
aDUP_CACCT_CLONE8     GTTCCTCACCCCGTGTCCCTCTTGGGGAAACAGGTTTCAGGAGCGATGGGTCTTGTAGCC    1029
aWT_GGTTC_BAC         GTTCCTCAC-CCGTGTCCCTCTTGGGGAAACAGGTTTCAGGAGCGACGGGTCTTGTAGCC    543
aWT_GGTTC_CLONE5      GTTCCTCAC-CCGTGTCCCTCTTGGGGAAACAGGTTTCAGGAGCGACGGGTCTTGTAGCC    543
                      ********* ************************************ *************

aDUP_CACCT_BAC        TGGGACAGCCAGGCCACCTGGGTGCAGCAATGCCTGAAGGCCTCCTGGCACCGAGACAGG    1320
aDUP_CACCT_CLONE8     TGGGACAGCCAGGCCACCTGGGTGCAGCAATGCCTGAAGGCCTCCTGGCACCGAGACAGG    1089
aWT_GGTTC_BAC         TGGGGCAGCCAGGCCACCTGGGTGCAGCTATGCCTGAAGGCCTCCTGGCACTGAGACAGG    603
aWT_GGTTC_CLONE5      TGGGGCAGCCAGGCCACCTGGGTGCAGCTATGCCTGAAGGCCTCCTGGCACTGAGACAGG    603
                      ****.***********************:********************** ********

aDUP_CACCT_BAC        GGCAGGAGCAGATCCCACCAGCGGGAAGGTGGTGGGTTCCAGTGCTGGGATCCACCAGCT    1380
aDUP_CACCT_CLONE8     GGCAGGAGCAGATCCCACCAGCGGGAAGGTGGTGGGTTCCAGTGCTGGGATCCACCAGCT    1149
aWT_GGTTC_BAC         GGCAGGAGCAGATCCCACCAGCGGGAAGGTGGTGCGTTCTGATGCTGGGATCCACCAGCT    663
aWT_GGTTC_CLONE5      GGCAGGAGCAGATCCCACCAGCGGGAAGGTGGTGCGTTCTGATGCTGGGATCCACCAGCT    663
                      ********************************** **** ..******************

aDUP_CACCT_BAC        GACAGGTGGAGCTGCCAGTCTCCAGTGCTCAGCCCTCAGCGGGGCCTGCCTGGCAGCCCC    1440
aDUP_CACCT_CLONE8     GACAGGTGGAGCTGCCAGTCTCCAGTGCTCAGCCCTCAGCGGGGCCTGCCTGGCAGCCCC    1209
aWT_GGTTC_BAC         GACAGGTGGAGCTGCGAGCCTCCAGTGCTCAGCCCTCGGCGGGGCCTGCCTGGCAGCCCC    723
aWT_GGTTC_CLONE5      GACAGGTGGAGCTGCGAGCCTCCAGTGCTCAGCCCTCGGCGGGGCCTGCCTGGCAGCCCC    723
                      *************** ** ******************.**********************

aDUP_CACCT_BAC        ACACACAGAGGGCATCGGGGTGGCGGGGGCACGTGTTACACGGGGGCCCTGGGTCTGAGT    1500
aDUP_CACCT_CLONE8     ACACACAGAGGGCATCGGGGTGGCGGGGGCACGTGTTACACGGGGGCCCTGGGTCTGAGT    1269
aWT_GGTTC_BAC         ACACACAGAGGGCATCGGGGTGGCGGGGGCACGTGTTACACGGGGGCCCTGGGTCTGAGT    783
aWT_GGTTC_CLONE5      ACACACAGAGGGCATCGGGGTGGCGGGGGCACGTGTTACACGGGGGCCCTGGGTCTGAGT    783
                      ************************************************************

aDUP_CACCT_BAC        CATCCACTTCCTCCGAGTCTGGATGGGAGGACCCAGCGCCCCTCCTCCGCCCCCTCCTGA    1560
aDUP_CACCT_CLONE8     CATCCACTTCCTCCGAGTCTGGATGGGAGGACCCAGCGCCCCTCCTCCGCCCCCTCCTGA    1329
aWT_GGTTC_BAC         CATCCACTTCCTCCGAGTCTGGATGGGAGGACCCAGCGCCCCTCCTCCGCCCCCTCCTGA    843
aWT_GGTTC_CLONE5      CATCCACTTCCTCCGAGTCTGGATGGGAGGACCCAGCGCCCCTCCTCCGCCCCCTCCTGA    843
                      ************************************************************

aDUP_CACCT_BAC        TCTGGAAGCATAAATGGGGAGGGGAGAGCCCACTGGGTAGAAGGAACAGGGAGCGGCCAG    1620
aDUP_CACCT_CLONE8     TCTGGAAGCATAAATGGGGAGGGGAGAGCCCACTGGGTAGAAGGAACAGGGAGCGGCCAG    1389
aWT_GGTTC_BAC         TCTGGAAGGATAAATGGGGAGGGGAGAGCCCGCTGGGTAGAAGGAACAGGGAGTGGCCAG    903
aWT_GGTTC_CLONE5      TCTGGAAGGATAAATGGGGAGGGGAGAGCCCGCTGGGTAGAAGGAACAGGGAGTGGCCAG    903
                      ******** **********************.********************* ******

aDUP_CACCT_BAC        GGTAAGTCCCCACTCTCAGAGACCCTGACATCAGCGTCACCTGGAGCAGAGTGGCCCAGC    1680
aDUP_CACCT_CLONE8     GGTAAGTCCCCACTCTCAGAGACCCTGACATCAGCGTCACCTGGAGCAGAGTGGCCCAGC    1449
aWT_GGTTC_BAC         GGTAAGTCCCTACTCTCAGAGACCCTGACATCAGCGTCACCTGGAGCAGAGTGGCCCAGC    963
aWT_GGTTC_CLONE5      GGTAAGTCCCTACTCCCAGAGACCCTGACATCAGCGTCACCTGGAGCAGAGTGGCCCAGC    963
                      ********** **** ********************************************

aDUP_CACCT_BAC        CTCAGACTCAGAGCACCAAGACCCAGGCCTGCAGGCCTGGACCCACCCCGGTCCCCCCGT    1740
aDUP_CACCT_CLONE8     CTCAGACTCAGAGCACCAAGACCCAGGCCTGCAGGCCTGGACCCACCCCGGTCCCCCCGT    1509
aWT_GGTTC_BAC         CTCAGACTCAGAGCACCAAGACCCAGGCCCGCAGGCCTGGACCCACCCCGGTCCCCCCGT    1023
aWT_GGTTC_CLONE5      CTCAGACTCAGAGCACCAAGACCCAGGCCCGCAGGCCTGGACCCACCCCGGTCCCCCCGT    1023
                      ***************************** ******************************

aDUP_CACCT_BAC        CCCAGCTCCATTCTTCACCCCACAATCTGTAGCCCCCAGCCCTGCCCTGTGAGGCCCGGC    1800
aDUP_CACCT_CLONE8     CCCAGCTCCATTCTTCACCCCACAATCTGTAGCCCCCAGCCCTGCCCTGTGAGGCCCGGC    1569
aWT_GGTTC_BAC         CCCAGCTCCATTCTTCACCCCACAATCTGTAGCCCCCAGCCCTGCCCTGTGAGGCCCGGC    1083
aWT_GGTTC_CLONE5      CCCAGCTCCATTCTTCACCCCACAATCTGTAGCCCCCAGCCCTGCCCTGTGAGGCCCGGC    1083
                      ************************************************************

aDUP_CACCT_BAC        CAGGCCCACGATGCTCCTCCTTGCTCCCCAGATG          1834
aDUP_CACCT_CLONE8     CAGGCCCACGATGCTC------------------          1585
aWT_GGTTC_BAC         CAGGCCCACGATGCTCCTCCTTGCTCCCCAGATG          1117
aWT_GGTTC_CLONE5      CAGGCCCACGATGCTCC-----------------          1100
                      ****************
```

**Figure S3. Multiple sequence alignment of representative cloned duplicated and wild-type α-tryptase promoters.**

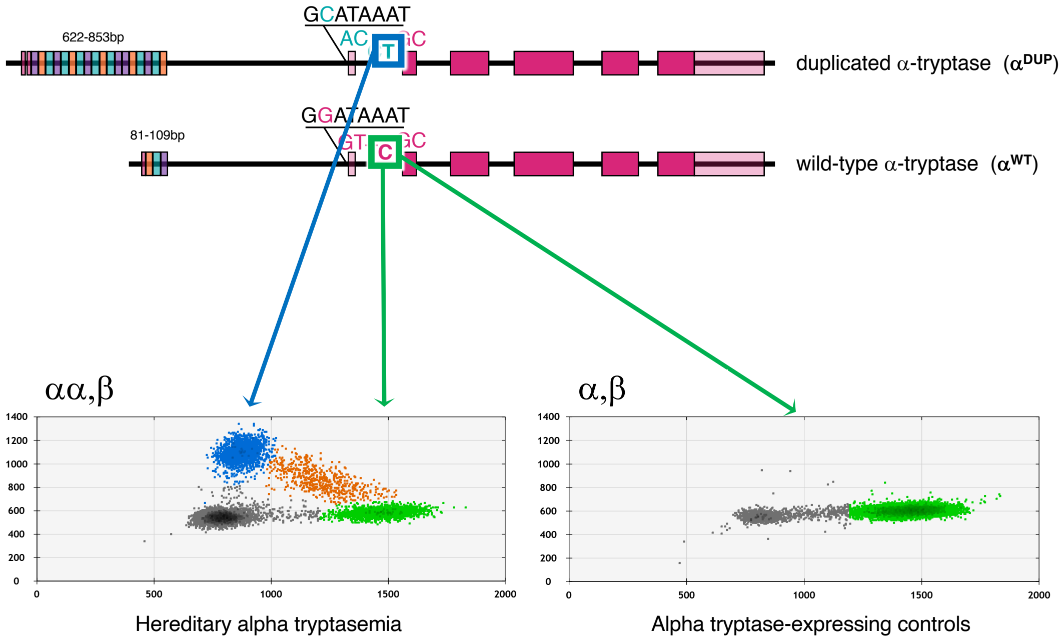

FWD: GGAGCAGAGTGGCCCAGCCTCAG
REV: ACGATGCCCACTCGCTGCAGGG (alpha-tryptase specific sequence in alpha$^{WT}$ and alpha$^{DUP}$)
PROBES: AGGCC**[C/T]**GCAGGCCTGGA

**Figure S4. Alpha-tryptase isoform-specific promoter assay.** (Top) Schematic of the two alpha-tryptase isoforms present at *TPSAB1* in a patient with hereditary alpha tryptasemia. In order to identify alleles containing the expanded promoter in linkage with duplicated alpha-tryptase ($\alpha^{DUP}$) in other samples, a reverse primer was designed to hybridize only to alpha-tryptase sequences, while the sequence complementary to the forward primer were present at all tryptase isoforms. Two probes were then designed and multiplexed in order to compete with one another to detect the single base pair change C>T, with the latter being present 5' of the first coding exon of $\alpha^{DUP}$ (primer/probe sequences at bottom). When only $\alpha^{WT}$ was present, fluorescence was detected in only one channel corresponding to the C-containing sequence (green, right middle). Whereas when $\alpha^{DUP}$ was present both a C linked to $\alpha^{WT}$ as well as the C>T were detected (blue, left middle).

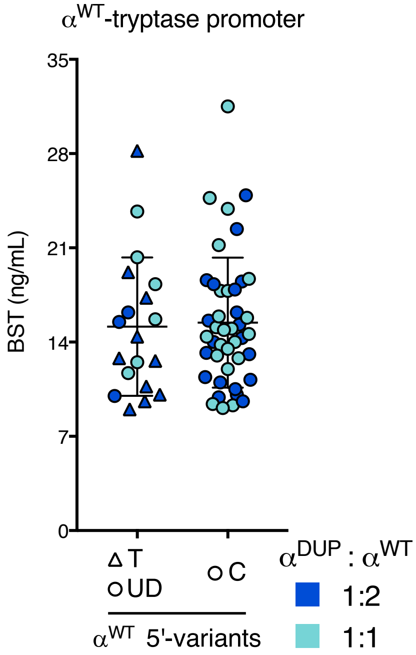

**Figure S5. Unidentified and C>T associated wild-type alpha-tryptase promoters do not affect basal serum tryptase.** Basal serum tryptase (BST) levels in patients with hereditary alpha tryptasemia separated by the promoter variant identified in linkage with wild-type alpha ($\alpha^{WT}$) on the allele harboring a duplication. While all $\alpha^{WT}$ sequences on non-duplicated alleles are linked to the major allele, some $\alpha^{WT}$ sequences on alleles with increased *TPSAB1* copy number were not. Individuals with the common major allele C are shown on the right, and individuals with undefined (UD, circles) or the minor allele T (triangles) are shown on the left. The ratio of duplicated alpha-tryptase ($\alpha^{DUP}$) to $\alpha^{WT}$ copy is indicated in blue (1:2) or cyan (1:1). Mann-Whitney.

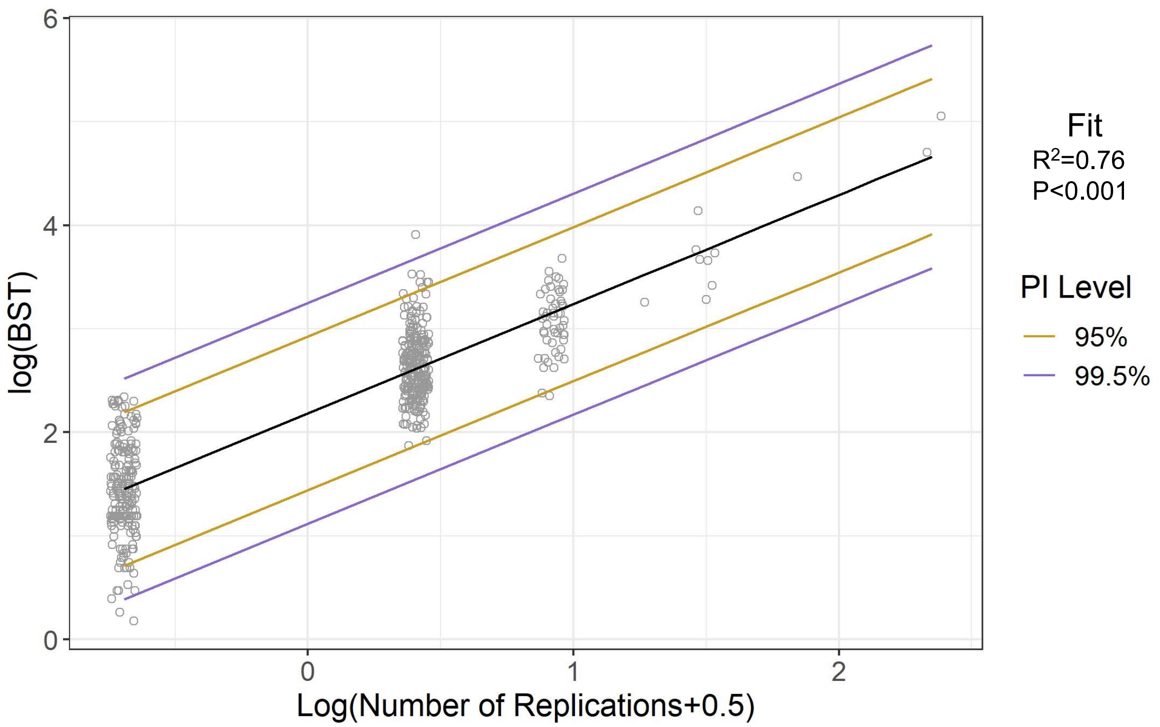

**Fig. S6. Log-transformed modeling of BST by *TPSAB1* copy number.**

**Table S1. Tryptase haplotype sequences.**

| | |
|---|---|
| CACCT | **C**ATAAATGGGGAGGGGAGAGCCC**A**CTGGGTAGAAGGAACA GGGAG**C**GGCCAGGGTAAGTCCC**C**ACTCTCAGAGACCCTGA CATCAGCGTCACCTGGAGCAGAGTGGCCCAGCCTCAGACT CAGAGCACCAAGACCCAGGCC**T** |
| GGTTC | **G**ATAAATGGGGAGGGGAGAGCCC**G**CTGGGTAGAAGGAACA GGGAG**T**GGCCAGGGTAAGTCCC**T**ACTCTCAGAGACCCTGA CATCAGCGTCACCTGGAGCAGAGTGGCCCAGCCTCAGACT CAGAGCACCAAGACCCAGGCC**C** |
| GACCC | **G**ATAAATGGGGAGGGGAGAGCCC**A**CTGGGTAGAAGGAACA GGGAG**C**GGCCAGGGTAAGTCCC**C**ACTCTCAGAGACCCTGA CATCAGCGTCACCTGGAGCAGAGTGGCCCAGCCTCAGACT CAGAGCACCAAGACCCAGGCC**C** |
| GACCT | **G**ATAAATGGGGAGGGGAGAGCCC**A**CTGGGTAGAAGGAACA GGGAG**C**GGCCAGGGTAAGTCCC**C**ACTCTCAGAGACCCTGA CATCAGCGTCACCTGGAGCAGAGTGGCCCAGCCTCAGACT CAGAGCACCAAGACCCAGGCC**T** |

**Table S2. 39-bp tryptase consensus sequences.**

| | |
|---|---|
| alpha$^{WT}$ | CCC**G**CTGGGTAGAAGGAACAGGGAG**T**GGCCAGGATGCTGA**GC** |
| alpha$^{DUP}$ | CCC**A**CTGGGTAGAAGGAACAGGGAG**C**GGCCAGGATGCTGA**GC** |
| beta | CCC**A**CTGGGTAGAAGGAACAGGGAG**C**GGCCAGGATGCTGA**AT** |

blue – 5’ UTR; black – first coding exon; red isoform-defining variants.

**Table S3. Tryptase primer/probe sequences.**

| | |
|---|---|
| $alpha^{TOTAL}$ | FWD: TGCTGCTGGCGCTGC<br>REV: GACTCTCAGGCTCACCTGC<br>PROBE: CTGCAGCAAGCGGGTATCGTC |
| $alpha^{WT}$ | FWD: CCCGCTGGGTAGAAGGAAC<br>REV: GACGATACCCGCTTGCTG<br>PROBE: TGGCCAGGATGCTGAGC |
| $alpha^{DUP}$ | FWD: CCCACTGGGTAGAAGGAAC<br>REV: GACGATACCCGCTTGCTG<br>PROBE: CGGCCAGGATGCTGAGC |
| beta | FWD: CCCACTGGGTAGAAGGAAC<br>REV: AACGATGCCCACTCGCTG<br>PROBE: CGGCCAGGATGCTGAAT |

### Table S4. Alpha-tryptase sequences and corresponding reference assemblies.

| Position (relative to TSS) | -30 | -8 | 36 | 44,45 | 177 | 309 | 333 | 508 | 644 | 647 | 657 | 661 |
|---|---|---|---|---|---|---|---|---|---|---|---|---|
| Alpha1; M30038.1; CHM1; alpha$^{WT}$ | G | T | G | GC | C | A | A | C | G | A | C | A |
| Alpha2; AF206665.1; BC051852.1; BC028059.1; LT733899.1; KJ892311.1; DQ894235.2; AY893004.1 | G | T | G | CG | T | G | A | C | C | G | A | C |
| AC238650.2; AF098328.1; AF206666.1; BC028059.1; BC051852.1; GM243385; NA12878 | G | T | C | CG | T | G | A | C | C | G | A | C |
| FJ931116.1 | G | T | G | CG | C | G | C | T | G | A | C | A |
| AC240106.3 | G | T | C | CG | T | G | A | C | G | A | C | A |
| alpha$^{DUP}$ | **A** | **C** | G | GC | C | A | A | C | G | A | C | A |

```
>Alpha1
cccgctgggtagaaggaacagggagtggccaggATGCTGAGCCTGCTGCTGCTGGCGCTGCCCGTCCTGGCGAGCCGCGCC
TACGCGGCCCCTGCCCCAGTCCAGGCCCTGCAGCAAGCGGGTATCGTCGGGGGTCAGGAGGCCCCCAGGAGCAAGTGGCCC
TGGCAGGTGAGCCTGAGAGTCCGCGACCGATACTGGATGCACTTCTGCGGGGGCTCCCTCATCCACCCCCAGTGGGTGCTG
ACCGCGGCGCACTGCCTGGGACCGGACGTCAAGGATCTGGCCACCCTCAGGGTGCAACTGCGGGAGCAGCACCTCTACTAC
CAGGACCAGCTGCTGCCAGTCAGCAGGATCATCGTGCACCCACAGTTCTACATCATCCAGACTGGAGCGGATATCGCCCTG
CTGGAGCTGGAGGAGCCCGTGAACATCTCCAGCCGCGTCCACACGGTCATGCTGCCCCCTGCCTCGGAGACCTTCCCCCCG
GGGATGCCGTGCTGGGTCACTGGCTGGGGCGATGTGGACAATGATGAGCCCCTCCCACCGCCATTTCCCCTGAAGCAGGTG
AAGGTCCCCATAATGGAAAACCACATTTGTGACGCAAAATACCACCTTGGCGCCTACACGGGAGACGACGTCCGCATCATC
CGTGACGACATGCTGTGTGCCGGGAACAGCCAGAGGGACTCCTGCAAGGGCGACTCTGGAGGGCCCCTGGTGTGCAAGGTG
AATGGCACCTGGCTACAGGCGGGCGTGGTCAGCTGGGACGAGGGCTGTGCCCAGCCCAACCGGCCTGGCATCTACACCCGT
GTCACCTACTACTTGGACTGGATCCACCACTATGTCCCCAAAAAGCCGTGA
```

**Table S5. Percentage of healthy individuals expressing specific α- and β-tryptase isoforms.**

| Transcript | Haplotype | Sample number (n = 4160) | % Total | % Tryptase positive |
|---|---|---|---|---|
| any tryptase | - | 863 | 20.7% | 100% |
| any α | ACAT | 741 | 17.8% | 85.9% |
| any β | NNGC | 314 | 7.5% | 36.4% |
| $\alpha^{DUP}$ | **ACGC** | **25** | **0.6%** | **2.9%** |
| $\alpha^{WT}$ | GTGC | 289 | 7.2% | 33.5% |

Data are from combined from 58 publicly available datasets; red – prevalence of hereditary alpha-tryptasemia

**Table S6. Candidate variants of undetermined significance identified in patients with BST >11.4ng/mL who did not have HαT.**

| Patient | Gene | Chr | Start | End | Ref | Var | AA change | PBMC | BMGran | BMMC |
|---|---|---|---|---|---|---|---|---|---|---|
| 1 | *TET2* | 4 | 105234636 | 105234636 | C | T | p.Q232X | 0.018923 | 0.2238426 | 0.1432696 |
| 1* | *FADS6* | 17 | 74893497 | 74893497 | C | CGGTTCCATG GGCTCCGTA | p.P33fs7X | 0.027044 | 0.1444073 | 0.1624679 |
| 1 | *GRB14* | 2 | 164508777 | 164508777 | G | GT | p.H297fs1X | 0 | 0.18850502 | 0.04259754 |
| 2 | *KMT2C* | 7 | 152247975 | 152247975 | G | A | p.T820I | 0.05543634 | 0.0711392 | 0.22914893 |
| 2* | *FADS6* | 17 | 74893497 | 74893497 | C | CGGTTCCATG GGCTCCGTA | p.P33fs7X | 0 | 0.1610098 | 0.08378218 |
| 2 | *KMT5A* | 12 | 123390764 | 123390764 | C | T | p.R49W | 0.02342595 | 0.06664452 | 0.26906613 |
| 2 | *MSH3* | 5 | 80654896 | 80654896 | G | C | p.A57P | 0.07519636 | 0.1615971 | 0.62264769 |
| 3 | *MAP2K1* | 15 | 66491063 | 66491063 | A | C | 3'UTR | 0 | 0.16156567 | 0.06649085 |
| 3 | *MAP2K1* | 15 | 66491059 | 66491059 | G | C | 3'UTR | 0 | 0.16472012 | 0.07177007 |
| 3 | *MAP2K1* | 15 | 66491055 | 66491055 | G | T | 3'UTR | 0 | 0.16472012 | 0.05015322 |
| 4 | *NOTCH2* | 1 | 120069404 | 120069404 | C | T | p.M1? | 0.14917127 | 0.20416667 | 0.20916334 |
| 4 | *RPL10* | X | 154400837 | 154400837 | C | T | p.R210W | 0 | 0.14081633 | 0.14257028 |
| 5† | *RUNX1* | 21 | 34792308 | 34792308 | A | G | p.S424P | 0.26548673 | 0.19277108 | 0.17142857 |
| 5† | *CEBPA* | 19 | 33301848 | 33301848 | GGGC | G | p.P189del | 0.09615385 | 0.01497006 | 0. 02076125 |

Chr - chromosome; Ref - reference; Var - variant; AA - amino acid; PBMC - peripheral blood mononuclear cell; BMGran - bone marrow granulocytes; BMMC - bone marrow mononuclear cells; *indicated a shared insertion present in patient 1 and 3; †identified variants were germline; patient 4 had idiopathic hypereosinophilic syndrome.
